# The Role of Sick-Quitter Bias in the Association Between Alcohol Use and Anxiety and Depression Symptoms: A Causal Analysis in a U.S. National Cohort

**DOI:** 10.64898/2026.07.25.26358931

**Authors:** Jenna Sanborn, Denis Nash, McKaylee Robertson, Angela Parcesepe, Zachary Shahn

## Abstract

**Background:** Observational studies frequently report a J- or U- shaped association between alcohol use and anxiety and depression, with moderate drinkers appearing to have lower risk than abstainers. However, this pattern may be driven in part by confounding due to health-related selection into abstinence or low-risk drinking (sick-quitter bias), whereby individuals reduce or stop drinking in response to declining health that is independently associated with worse mental health outcomes. We aimed to evaluate whether this association reflects a causal effect or residual confounding driven by sick-quitter bias.

**Methods:** We analyzed data from 4,673 participants in the CHASING COVID Cohort, a U.S. longitudinal study with repeated measures from September 2021 through December 2023. Alcohol use was assessed at five timepoints using the AUDIT-C and categorized as abstinent, low-risk, moderate-risk, or high/severe-risk. Using a target trial emulation framework, we estimated the effects of sustained alcohol use strategies on anxiety and depression symptom severity (GAD-7 and PHQ-8) at follow-up. Artificial censoring and stabilized inverse probability weighting were applied to account for time-varying confounding and loss to follow-up. To assess the role of sick-quitter bias, we compared the abstinent versus moderate-risk contrast across strata of health status at time zero.

**Results:** In the full sample, a J-shaped pattern was observed, with moderate-risk drinkers having the lowest predicted symptom scores. The abstinent versus moderate-risk symptom score contrast was 1.38 for depression (PHQ-8: 95% CI: 0.65, 2.14), and 0.95 for anxiety (GAD-7: 95% CI 0.24, 1.65). Among participants without underlying conditions or poor self-rated health, this pattern attenuated, with contrasts near zero for both outcomes. In contrast, the J-shaped pattern was amplified among participants with poorer baseline health, with abstinent versus moderate-risk contrasts of 3.31 for depression (95% CI: 2.14, 4.32) and 2.83 for anxiety (95% CI: 1.61, 3.86). High-risk drinking was consistently associated with higher symptom scores across all analyses.

**Conclusions:** The J-shaped pattern between alcohol use and anxiety and depression symptoms observed in the full cohort was attenuated among participants without poorer baseline health, consistent with sick-quitter bias. Findings underscore the importance of addressing confounding due to declining health in observational alcohol research and caution against interpreting moderate drinking as beneficial for symptoms of anxiety and depression.

## Introduction

Depression and anxiety are highly prevalent in the United States (US), affecting more than one in five adults and ranking among the leading contributors to global disability.^1–3^ In 2021, an estimated 48 million adults experienced anxiety, while 21 million had a major depressive episode.^4,5^ These disorders frequently co-occur^6,7^ and are associated with elevated risks of chronic disease and premature mortality.^8–16^ Efforts to reduce the burden of anxiety and depression require attention to modifiable risk factors. Clarifying the role of alcohol use is especially important given its ubiquity and established links to poor mental health. In 2024, over 174 million US adults consumed alcohol and more than 28 million met criteria for alcohol use disorder (AUD).^17,18^ AUD is strongly linked to depression and anxiety, with evidence of a bidirectional causal relationship.^19–24^ However, the relationship between moderate alcohol use and anxiety and depression remains unclear. Most studies, including a recent meta-analysis, have observed a J- or U- shaped pattern, in which moderate drinkers appear to have lower risk than abstainers.^25–34^ Similar patterns have been reported in studies of cardiovascular disease and mortality, where apparent benefits of moderate drinking were later attributed to bias rather than causal protection.^35–39^

Several methodological issues complicate causal inference when alcohol is the exposure. Most prior studies examining the impact of alcohol on anxiety and depression have relied on cross-sectional data or single baseline measures of alcohol use,^7,20,25,26,29,40–47^ limiting the ability to establish temporality and failing to account for changes in alcohol consumption over time.^48,49^ Moderate drinkers have also been shown to systematically differ from abstainers in sociodemographic, behavioral, and health-related characteristics, leading to residual confounding when these factors are not accounted for.^48,50–53^ Importantly, individuals who reduce or stop drinking due to declining health (“sick-quitters”) may have worse mental health outcomes independent of alcohol use, thereby confounding associations with abstinence and low-risk drinking and inflating estimates that make moderate drinkers appear more protected.^48,51,54,55^ Furthermore, most longitudinal studies have overlooked the dynamic nature of alcohol consumption, leading to time-varying treatment-confounder feedback, where prior alcohol use influences later covariates that in turn affect subsequent drinking and mental health outcomes.^56–58^ G-methods are a class of causal inference methods designed to handle time-varying confounding, but have rarely been applied in this context. In the two longitudinal studies that that have employed g-methods to examine alcohol use and depression, moderate drinking was associated with lower risk of depression compared with abstinence.^31,34^ However, neither accounted for sick-quitter bias, leaving uncertainty about whether lower symptom burden among moderate-drinkers relative to abstainers reflects a causal effect of moderate drinking or residual confounding.

These challenges raise the concern that lower anxiety and depression symptom burden among moderate-risk drinkers relative to abstainers may reflect confounding due to health-related selection into abstinence or low-risk drinking (sick-quitter bias) rather than a true protective effect of moderate drinking. However, sick-quitter bias and time-varying confounding have received limited attention in studies examining alcohol’s impact on mental health. As a result, misinterpretations of apparent protective effects of moderate alcohol use have misguided public health messaging.^59^ Several reviews have therefore called for research that more rigorously evaluates causal mechanisms and applies advanced causal inference methods to strengthen the evidence base for guidelines and interventions.^23,25,34,60–62^

To evaluate whether lower anxiety and depression symptom severity under moderate-risk drinking relative to abstinence reflects a causal effect or residual confounding due to sick-quitter bias, we analyzed data from the CHASING COVID Cohort, a large U.S. longitudinal study with repeated measures of alcohol use, anxiety and depression symptoms, and time-varying covariates. Using artificial censoring with inverse probability weighting to estimate the g-formula, we estimated mean symptom scores under sustained alcohol use strategies.^63,64^ We used the contrast in predicted symptoms between sustained abstinent and sustained moderate-risk drinking as the primary measure of the lower symptom burden observed among moderate-risk drinkers. To assess the role of sick-quitter bias, we compared this contrast across strata of health status at time zero, hypothesizing that the it would attenuate among participants without underlying conditions or poor self-rated health and amplify among those with poorer baseline health.

## Methods

### Study Population & Analytic Sample

The CHASING COVID Cohort is a national, community-based prospective study of 5,798 adults (aged ≥18) recruited in 2020 and followed approximately every three months through 2023. Participants resided in all 50 U.S. states, Washington D.C., Puerto Rico, and Guam and was socio-demographically diverse. Detailed recruitment methods and follow-up procedures have been published,^65^ and study materials are publicly available.^66^ For this analysis, the September 2021 assessment was defined as time zero, corresponding to the post-vaccine period when the acute phase of the pandemic had stabilized. Among 5,798 cohort participants, 4,673 with valid time zero alcohol consumption data as well as sex information, required for Alcohol Use Disorders Identification Test-Consumption (AUDIT-C) classification, were included in the analytic sample (Figure 1).

**Figure 1.**
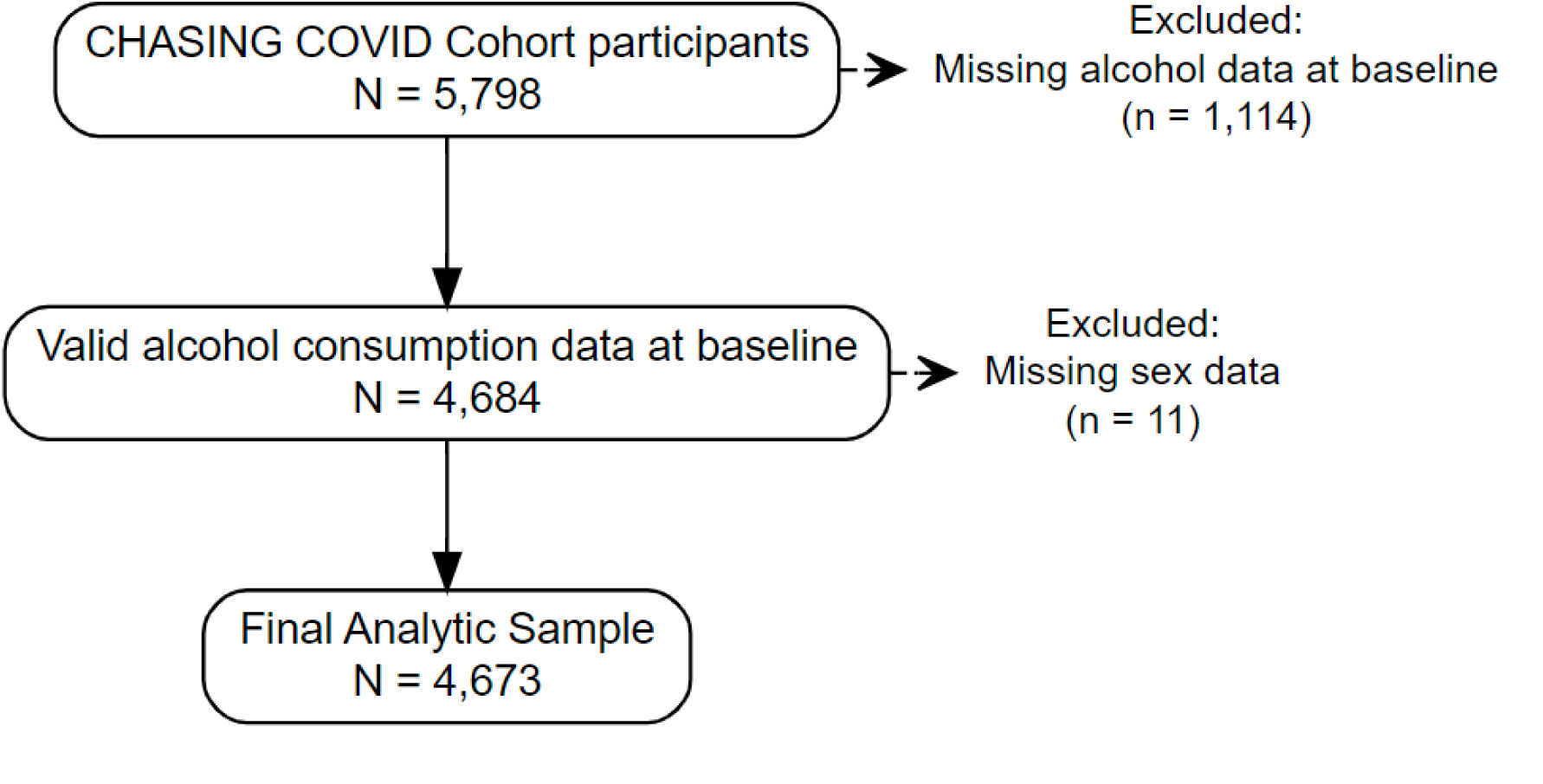
Flow Diagram of Participant Inclusion and Exclusions.

### Research Ethics

The study protocol was approved by the Institutional Review Board at the City University of New York (CUNY) Graduate School for Public Health and Health Policy. Participant consent was obtained at baseline and at periodic follow-up assessments. Participants could voluntarily discontinue participation at any time.

### Alcohol Consumption (Exposure)

Alcohol use was assessed at five timepoints (September 2021, March 2022, October 2022, April 2023, and September 2023) using the 3-item AUDIT-C, a validated screening tool to identify unhealthy alcohol use. The scale sums three items on drinking frequency, typical quantity, and frequency of heavy drinking, with scores ranging from 0 (no use) to 12.^67^ We categorized consumption into abstinence, low risk, moderate risk, and high/severe risk groups, using sex-specific cutoff scores based on guidelines adapted from the U.S. Department of Veterans Affairs: abstinence (score = 0 for both sexes), low risk (scores 1-2 for females, 1-3 for males), moderate risk (scores 3-5 for females, 4-5 for males), and high/severe risk (score ≥6 for both sexes).^68^

### Anxiety and Depression Symptom Severity (Outcomes)

Symptoms of anxiety and depression were measured at each timepoint using the validated Generalized Anxiety Disorder-7 (GAD-7; score range 0-21) and Patient Health Questionnaire-8 (PHQ-8; score range 0-24).^69–73^ Outcomes were analyzed separately as continuous measures of symptom severity at the final assessment in December 2023.

### Covariates

We adjusted for time-fixed and time-varying covariates selected based on literature, subject-matter knowledge, and directed acyclic graphs. Time-fixed covariates included sociodemographic characteristics (age, gender, race/ethnicity, education, income, body-mass index, daily smoking, household size, residential area type), mean AUDIT-C scores measured prior to time zero, neighborhood community, life satisfaction and spirituality/religion. Time-varying covariates included prior anxiety and depression symptoms, physical activity, material hardship, healthcare access, disability status, number of diagnosed chronic conditions, diagnosed mental health conditions, mental health treatment, self-reported health status, social engagement, substance use, relationship status, recovery from drugs or alcohol, and COVID-19 vaccination status. To preserve temporal ordering, time-varying covariates were drawn from prior assessments. Full variable definitions are provided in Supplementary Table S1.

### Statistical analysis

We used a target trial emulation framework to estimate the effect of sustained alcohol use strategies on symptoms of anxiety and depression. The target trial is conceptualized as a hypothetical randomized trial in which participants are “assigned” at time zero (September 2021) to sustained alcohol use strategies defined by their observed alcohol use category at time zero and followed through December 2023. Because treatment assignment is not randomized in the observed data, identification relies on the assumption of conditional exchangeability given measured baseline and time-varying covariates.

We evaluated four sustained alcohol-use strategies-abstinent, low-risk, moderate-risk, and high/severe-risk drinking-defined by consistent adherence to the same AUDIT-C category across all assessment waves. These strategies were emulated using artificial censoring, whereby participants were censored at the first assessment at which their observed alcohol use deviated from the assigned strategy. Stabilized inverse probability weights were used to account for time-varying confounding and censoring arising from both the artificial censoring and loss to follow-up, with weights estimated from models including baseline and time-varying covariates. Intuitively, this approach creates a weighted pseudo-population in which adherence to each strategy is independent of measured covariates, approximating the exchangeability that would be achieved under randomization. We then estimated weighted mean PHQ-8 and GAD-7 scores at the final assessment under each sustained strategy.

Because higher symptom scores under sustained abstinence relative to sustained moderate-risk drinking would indicate J- or U-shaped patterning in predicted symptom severity, we computed contrasts comparing abstinence with moderate-risk drinking. Confidence intervals were obtained using nonparametric bootstrap resampling (B = 1000), with the full estimation procedure repeated in each replicate. Additional details on model specification, weight construction, and diagnostics are provided in the Supplementary Materials.

All analyses were conducted in R (version 4.5.2)

### Subgroup Analyses

To illustrate the potential for sick-quitter bias, we present a directed acyclic graph depicting prior illness, past problem drinking, and their shared causes may lead individuals to reduce or stop drinking because of declining health while also independently increasing subsequent anxiety and depression symptoms, creating a backdoor path between abstinence/low-risk drinking and mental health outcomes through being “sick enough to quit” (Figure 2). This pathway is difficult to fully address when health-related drinking reduction or cessation occurred before cohort enrollment, because we did not directly measure prior sick-quitting nor observe the health, mental health, and drinking histories that could otherwise serve as proxies for that selection process. Although repeated measures from April 2020 onward allowed us to adjust for observed health, mental health, and drinking-related proxies of sick-quitting before and during follow-up, more remote histories remain incompletely captured. We therefore used health status as a proxy for susceptibility to this source of bias: participants without underlying conditions or poor self-rated health were less likely to include individuals who reduced or stopped drinking because of declining health prior to enrollment, whereas among participants with poorer health, prior sick quitters may be present alongside non-sick quitters, leaving greater potential for residual confounding.

**Figure 2.**
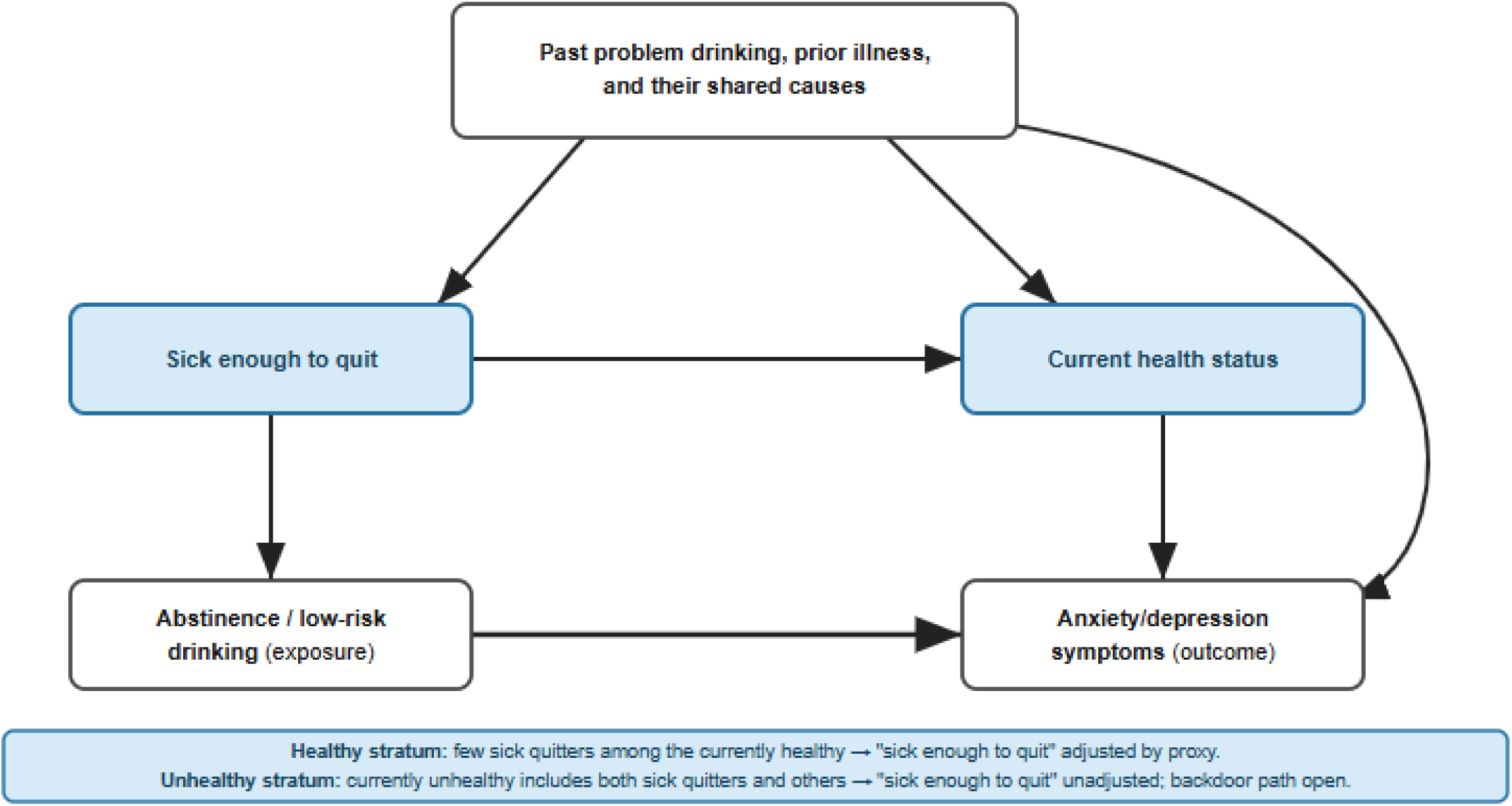
Causal diagram illustrating the backdoor path linking abstinence to anxiety/depression symptoms via confounding due to health-related selection into abstinence (sick-quitter bias). Causal diagram illustrating how health-related selection (sick-quitter bias) confounds the association between abstinence/low-risk drinking and anxiety/depression symptoms. Past problem drinking, prior illness, and their shared causes can lead individuals to quit drinking and independently worsen mental health, creating a backdoor path through “sick enough to quit” Restricting to currently healthy individuals effectively excludes sick quitters, so this node is adjusted by proxy. Among those with underlying conditions or poor health, sick quitters and non-sick quitters coexist, leaving the backdoor path open and confounding unresolved.

To assess whether elevated symptom scores under sustained abstinence relative to sustained moderate-risk drinking may reflect this selection mechanism, we stratified analyses by health status at time zero. We compared participants without chronic conditions or poor self-rated health (N=2,813), in whom such selection is less plausible, to those with underlying conditions or poor self-rated health (N=1,860), in whom it is more likely. Chronic conditions were defined based on self-reported diagnoses (including diabetes, high blood pressure, chronic lung disease or COPD, heart attack, angina, kidney disease, immunocompromised condition, HIV, or current asthma) at enrollment. Poor general health was defined as any report of “poor” general health across four assessments conducted from July 2020-September 2021.

### Sensitivity Analysis

As a sensitivity analysis, we repeated the analysis using an alternative lag structure for time-varying covariates (t0 and t-1 instead of t-1 and t-2) to assess robustness to covariate timing assumptions. Additional details are provided in the Supplement.

### Effect modification by sex

Given known differences in alcohol use and mental health across sexes, we assessed effect modification by sex using interaction terms between alcohol use category and sex in weighted regression models, applying the same stabilized inverse probability weights as in the primary analysis.^74–76^

## Results

### Sample characteristics

Table 1 shows participant characteristics at time zero. About one-third (30.3%) of participants were abstinent (n=1,418), 37.4% were low-risk drinkers (n=1,748), 22.9% were moderate-risk drinkers (n=1,069), and 9.4% were high/severe-risk drinkers (n=438). Participant characteristics varied markedly by alcohol category. Compared with all other categories, high/severe risk drinkers were younger, more often male, more likely to reside in urban areas, and more likely to smoke daily. They also shared several indicators of socioeconomic vulnerability with abstainers, including higher rates of housing insecurity, food insecurity, lower educational attainment, higher unemployment, more frequent use of government food assistance, and lower physical activity compared with low- and moderate-risk drinkers. Identification as being in recovery from drugs and/or alcohol was more common among both abstinent and high/severe groups than among low- and moderate-risk groups.

**Table 1.**
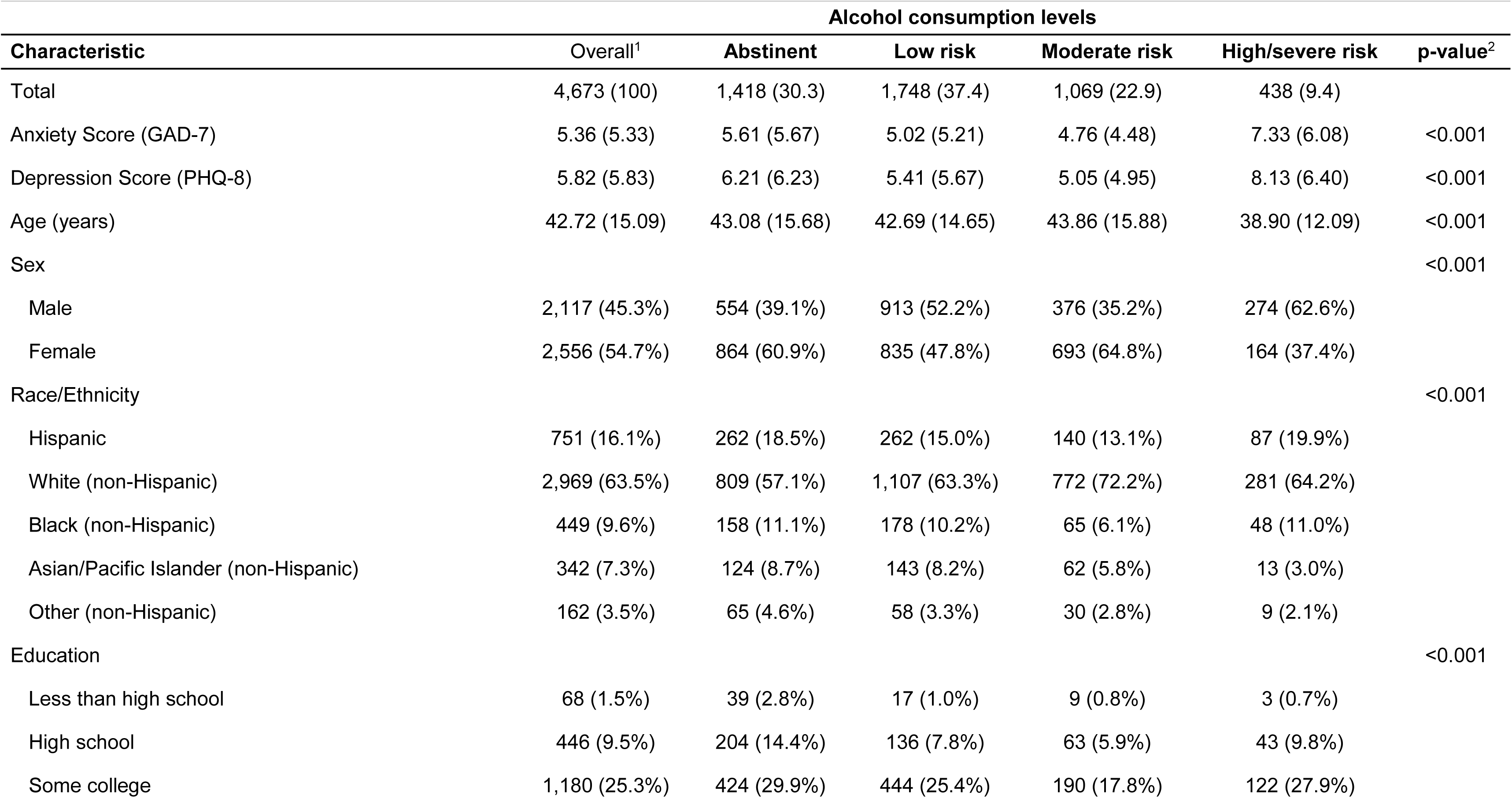
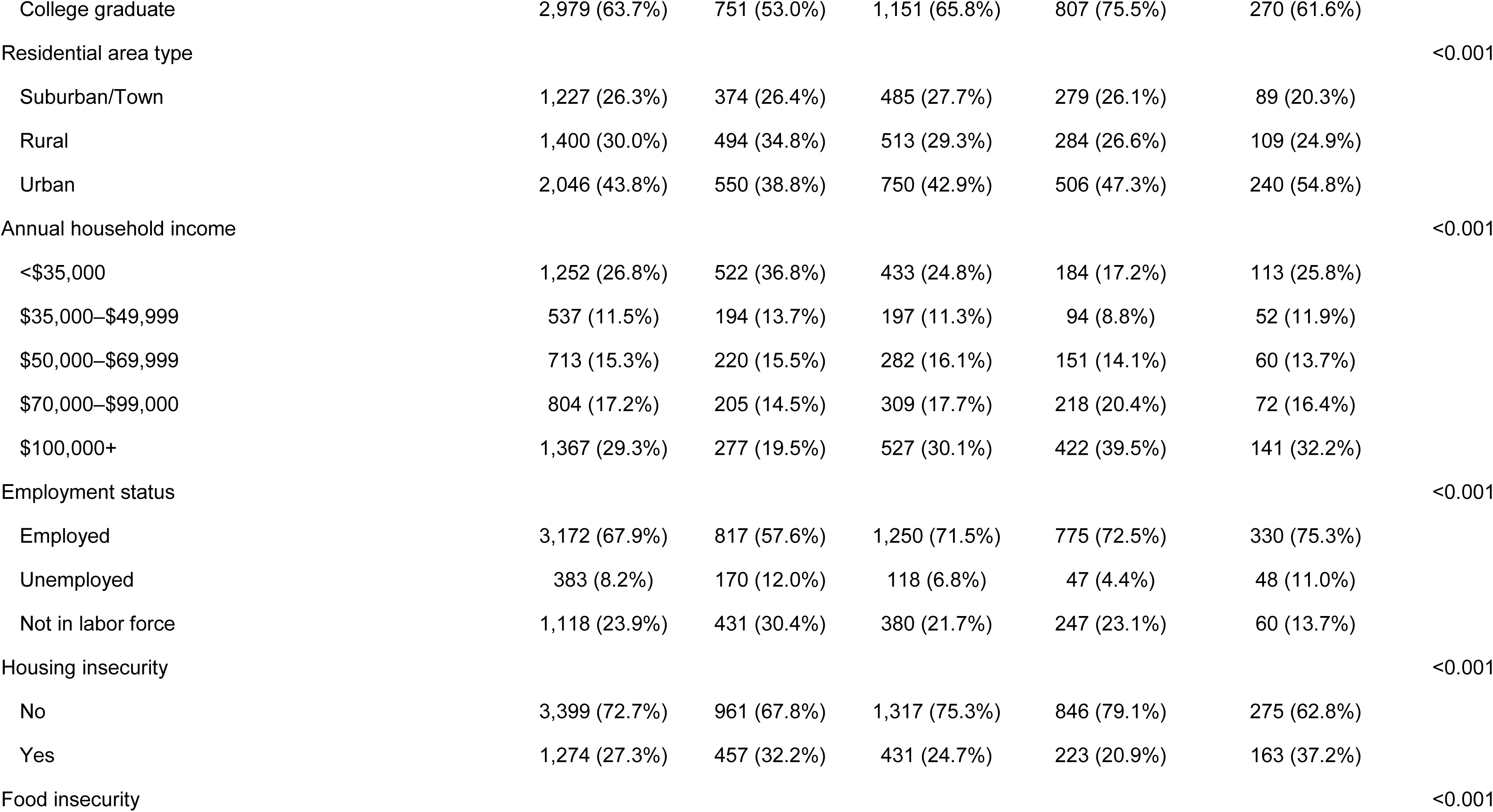
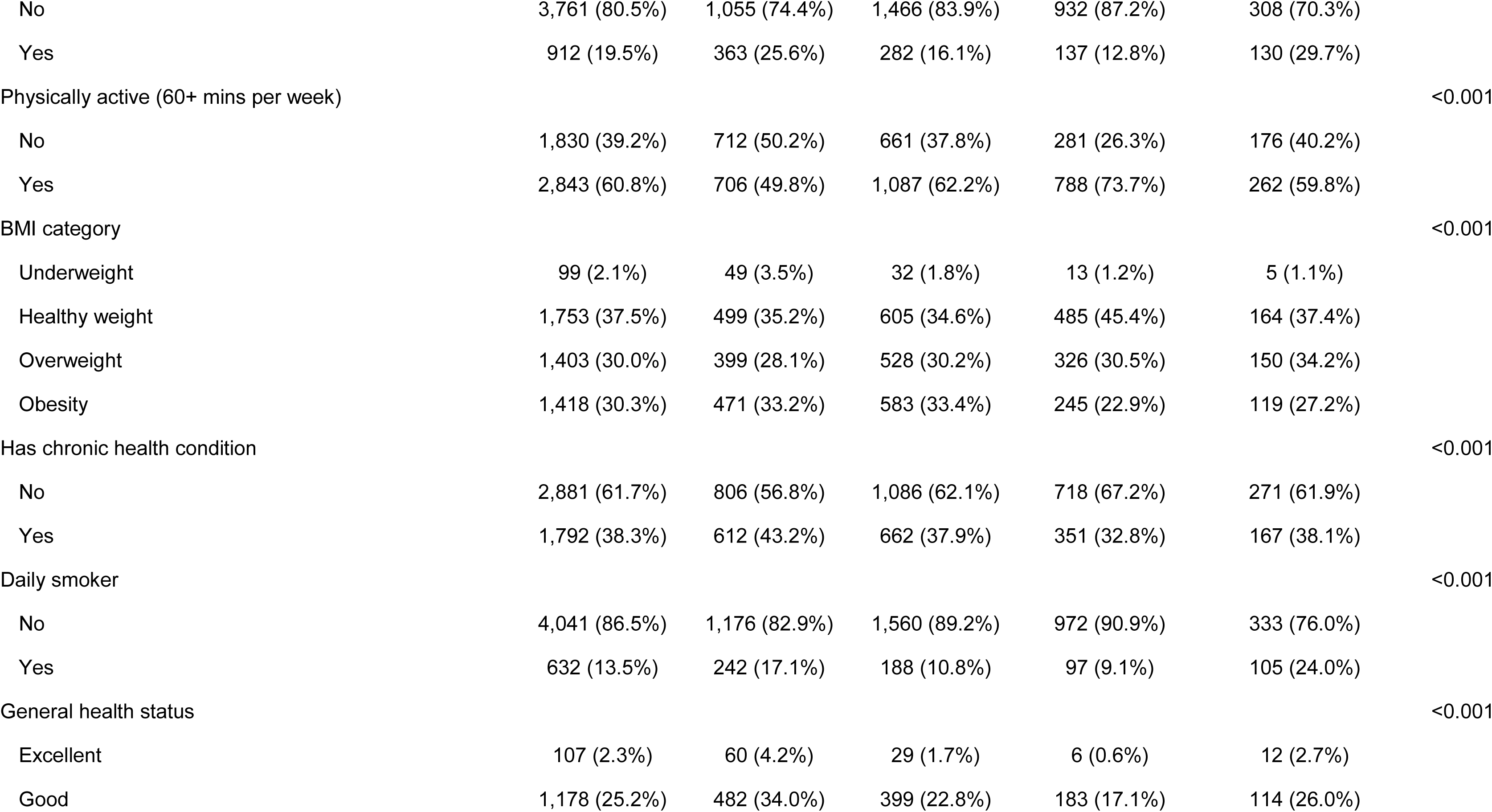
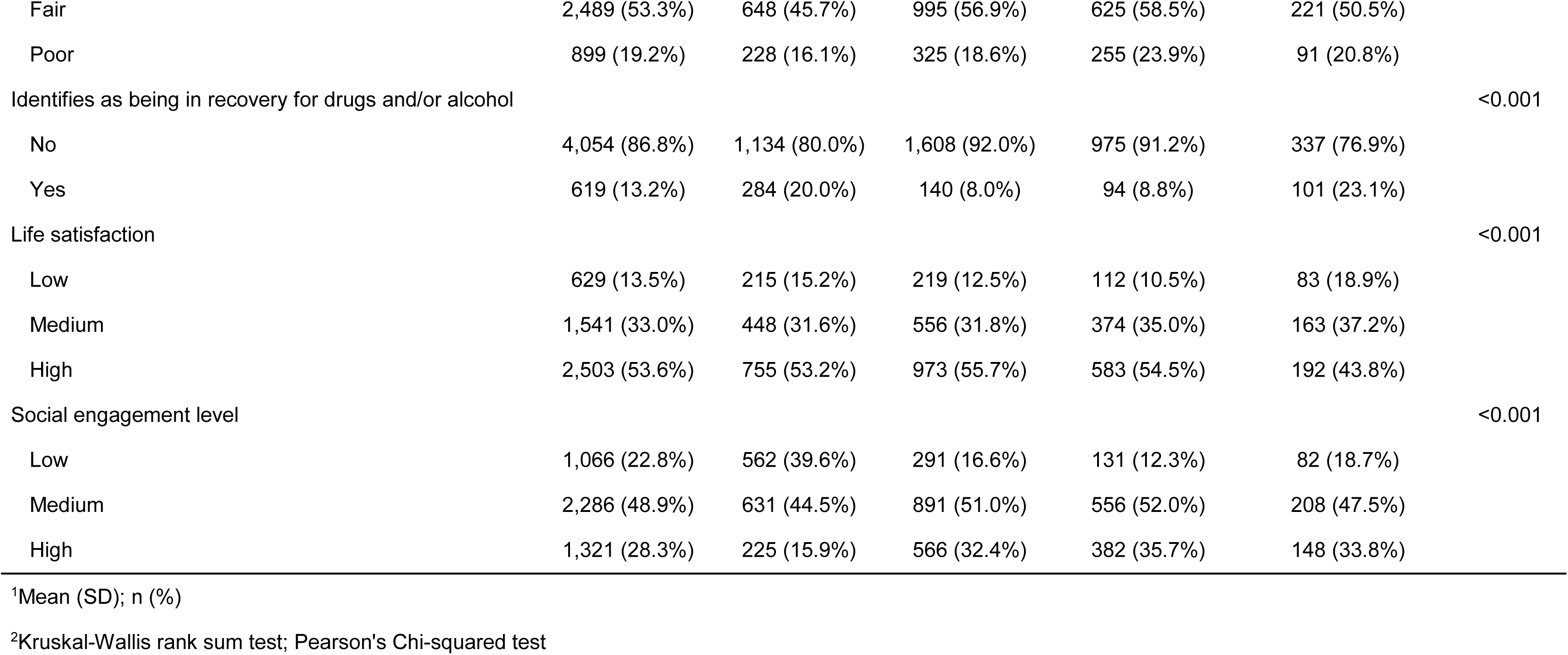
Select participant characteristics by baseline alcohol consumption, CHASING COVID Cohort, September 2021 (N=4,673)

Compared with all other drinking groups, abstainers were more likely to reside in rural areas, to report lower household income, to be out of the labor force, and to have a chronic health condition. They were also less likely to be White non-Hispanic, college graduates, or physically active, and had the lowest levels of social engagement of any group. Moderate-risk drinkers presented a notably different profile. The majority were female (64.8%) and White non-Hispanic (72.2%), and they were more likely than all other groups to be college educated, employed, and high-income (≥$100,000 per year). They also reported the highest levels of social engagement and physical activity, and the lowest prevalence of food and housing insecurity of any alcohol category.

Characteristics across the three analytic subsamples are presented in Supplementary Table S2; as expected, participants with underlying conditions or poor self-rated health were older and showed greater socioeconomic vulnerability compared with those without.

### Primary analyses

Predicted mean symptom scores under sustained alcohol use strategies estimated using artificial censoring and inverse probability weighting are summarized in Table 2. In the full sample (N = 4,673), a J-shaped association was observed for both outcomes. For depression (PHQ-8; Figure 3A), the predicted mean scores were 4.72 (95% CI: 4.33, 5.16) for sustained abstinence, 4.21 (3.66, 5.03) for low risk, 3.34 (2.78, 3.96) for moderate risk, and 6.11 (5.00, 7.73) for high risk, yielding an abstinent-moderate contrast of 1.38 (0.65, 2.14). For anxiety (GAD-7; Figure 4A), predicted means were 3.95 (3.56, 4.33), 3.40 (3.02, 4.01), 3.00 (2.46, 3.57), and 5.42 (4.35, 6.90) respectively, with an abstinent-moderate contrast of 0.95 (0.24, 1.65).

**Figure 3.**
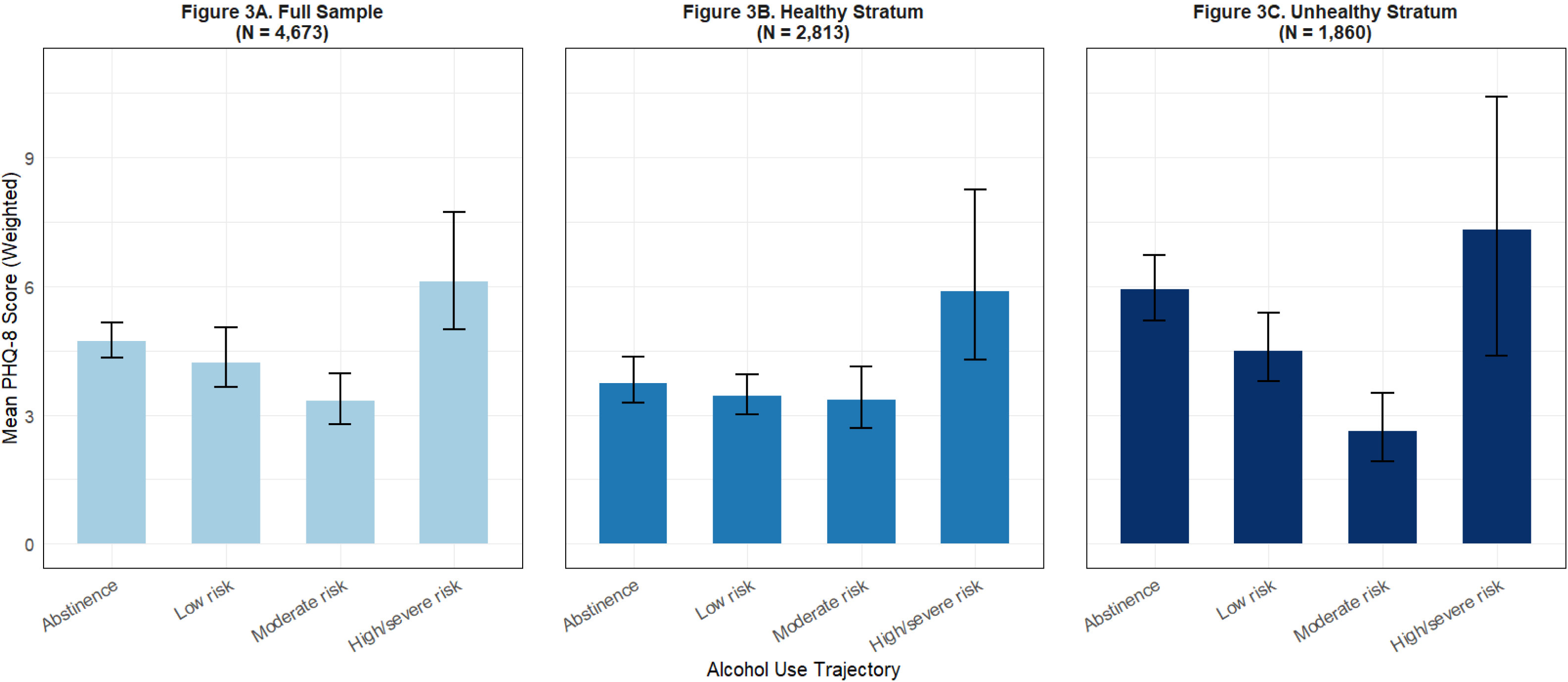
Predicted Mean PHQ-8 Scores Under Sustained Alcohol Use Trajectories. Predicted mean depression (PHQ-8) and anxiety (GAD-7) symptom severity in December 2023 under sustained alcohol use trajectories from September 2021 to September 2023, estimated using inverse probability weighting with artificial censoring. Panel A displays results for the full sample (N = 4,673), Panel B displays results for the healthy stratum, excluding participants with chronic conditions or poor self-rated health (N = 2,813), and Panel C displays results for the unhealthy stratum, restricted to participants with chronic conditions or poor self-rated health (N = 1,860). In the full sample, a J-shaped pattern was observed for both outcomes, with higher symptom scores among abstainers and high/severe-risk drinkers relative to low- and moderate-risk groups. This pattern was attenuated in the healthy stratum and amplified in the unhealthy stratum, consistent with sick-quitter bias and health-related selection into abstinence. Error bars represent 95% confidence intervals based on 1,000 nonparametric bootstrap samples.

**Figure 4.**
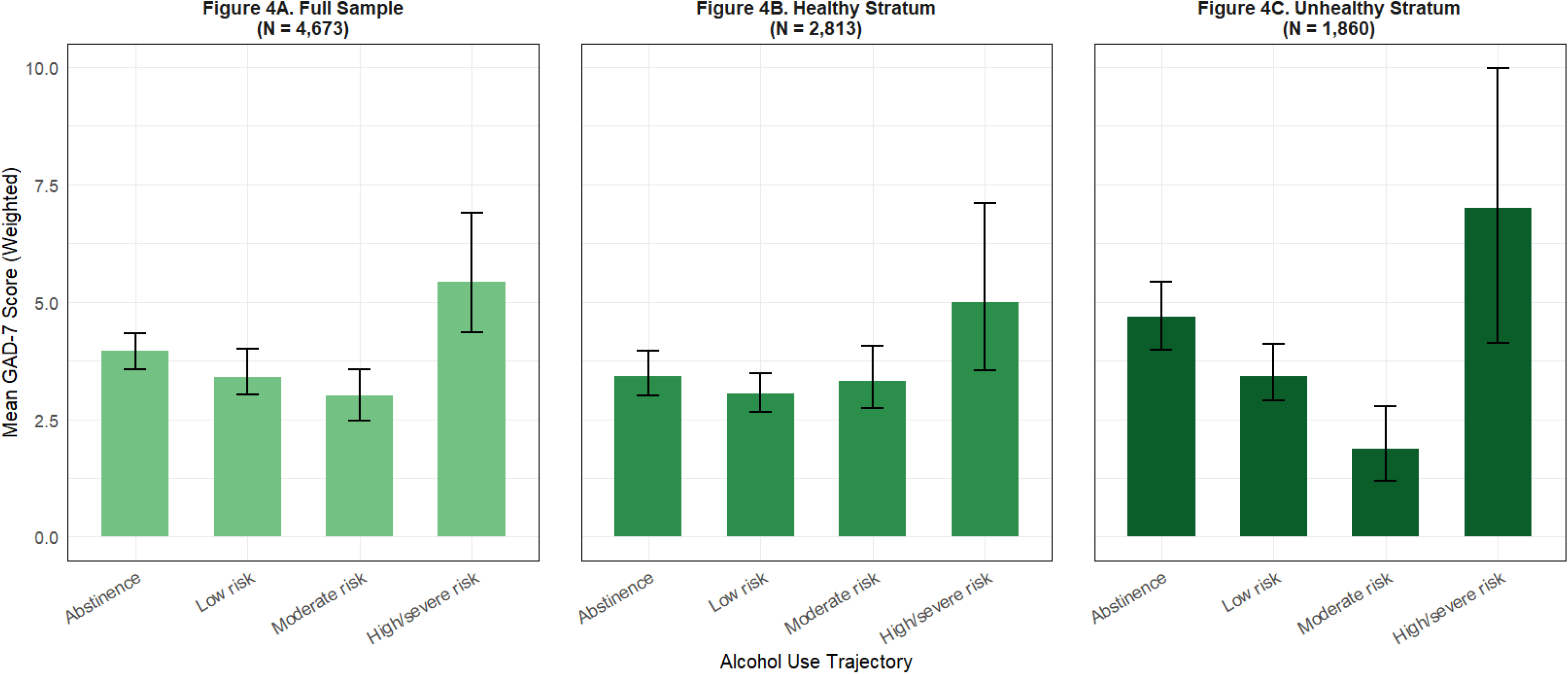
Predicted Mean GAD-7 Scores Under Sustained Alcohol Use Trajectories. Predicted mean depression (PHQ-8) and anxiety (GAD-7) symptom severity in December 2023 under sustained alcohol use trajectories from September 2021 to September 2023, estimated using inverse probability weighting with artificial censoring. Panel A displays results for the full sample (N = 4,673), Panel B displays results for the healthy stratum, excluding participants with chronic conditions or poor self-rated health (N = 2,813), and Panel C displays results for the unhealthy stratum, restricted to participants with chronic conditions or poor self-rated health (N = 1,860). In the full sample, a J-shaped pattern was observed for both outcomes, with higher symptom scores among abstainers and high/severe-risk drinkers relative to low- and moderate-risk groups. This pattern was attenuated in the healthy stratum and amplified in the unhealthy stratum, consistent with sick-quitter bias and health-related selection into abstinence. Error bars represent 95% confidence intervals based on 1,000 nonparametric bootstrap samples.

**Table 2.** Predicted Mean GAD-7 and PHQ-8 scores by Alcohol Use Trajectory. (Point Estimates and 95% Confidence Intervals)

|  |  |  |  |  |  |  |
| --- | --- | --- | --- | --- | --- | --- |
| Strategy | GAD-7 Full Cohort<br>(N=4,673) | GAD-7 Healthy<br>Stratum (N=2,813) | GAD-7 Unhealthy<br>Stratum (N=1,860) | PHQ-8 Full Cohort<br>(N=4,673) | PHQ-8 Healthy<br>Stratum (N=2,813) | Unhealthy Stratum<br>(N=1,860) |
| Abstinent | 3.95 [3.56, 4.33] | 3.43 [3.01, 3.96] | 4.69 [3.99, 5.43] | 4.72 [4.33, 5.16] | 3.74 [3.29, 4.36] | 5.93 [5.19, 6.72] |
| Low risk | 3.40 [3.02, 4.01] | 3.04 [2.65, 3.49] | 3.42 [2.90, 4.11] | 4.21 [3.66, 5.03] | 3.44 [3.02, 3.95] | 4.50 [3.79, 5.39] |
| Moderate risk | 3.00 [2.46, 3.57] | 3.32 [2.73, 4.06] | 1.86 [1.19, 2.78] | 3.34 [2.78, 3.96] | 3.36 [2.70, 4.13] | 2.62 [1.91, 3.52] |
| High risk | 5.42 [4.35, 6.90] | 5.00 [3.54, 7.10] | 7.00 [4.12, 9.98] | 6.11 [5.00, 7.73] | 5.87 [4.29, 8.25] | 7.32 [4.37, 10.4] |
| Abstinent –<br>Moderate contrast | 0.95 [0.24, 1.65] | 0.11 [-0.73, 0.89] | 2.83 [1.61, 3.86] | 1.38 [0.65, 2.14] | 0.38 [-0.49, 1.25] | 3.31 [2.14, 4.32] |

Healthy stratum excludes participants with any underlying chronic condition or poor self-rated health at time zero. Unhealthy stratum is restricted to participants with any underlying chronic condition or poor self-rated health at time zero. Underlying chronic conditions included diabetes, high blood pressure, chronic lung disease or COPD, heart disease, kidney disease, immunocompromising condition, HIV, chronic liver disease, and current asthma.

In the healthier subsample without underlying conditions or poor self-rated health (N = 2,813), the J-shaped association was attenuated (Table 2). For PHQ-8 (Figure 3B), predicted means were 3.74 (95% CI: 3.29, 4.36) for abstinence, 3.44 (3.02, 3.95) for low risk, 3.36 (2.70, 4.13) for moderate risk, and 5.79 (4.27, 7.77) for high risk; the abstinent-moderate contrast was 0.38 (−0.49, 1.25). For GAD-7 (Figure 4B), predicted means were 3.43 (3.01, 3.96), 3.04 (2.65, 3.49), 3.32 (2.73, 4.06), and 5.00 (3.54, 7.10), respectively; the abstinent-moderate contrast was 0.11 (−0.73, 0.89).

In the subsample with underlying conditions or poor self-rated health (N=1,860), the J-shaped pattern was substantially amplified (Table 2). For PHQ-8, predicted means were 5.93 (5.19, 6.72) for abstinence, 4.50 (3.79, 5.39) for low risk, 2.62 (1.91, 3.52) for moderate risk, and 7.32 (4.37, 10.40) for high risk; the abstinent-moderate contrast was 3.31 (2.14, 4.32), more than double that observed in the full sample and nearly nine times that observed in the subsample without underlying conditions or poor self-rated health (Figure 3C). For GAD-7, predicted means were 4.69 (3.99, 5.43), 3.42 (2.90, 4.11), 1.86 (1.19, 2.78), and 7.00 (4.12, 9.98), respectively; the abstinent-moderate contrast was 2.83 (1.61, 3.86) (Figure 4C).

Within-strategy comparisons showed that restricting to participants without underlying conditions or poor self-rated health attenuated predicted PHQ-8 scores for abstainers by approximately one point (4.72 [4.33, 5.16] vs. 3.74 [3.29, 4.36]) and for low-risk drinkers by approximately 0.8 points (4.21 [3.66, 5.03] vs. 3.44 [3.02, 3.95]), while predicted scores for moderate-risk drinkers were virtually unchanged (3.34 [2.78, 3.96] vs. 3.36 [2.70, 4.13]), consistent with sick-quitter bias rather than a genuine protective effect of moderate drinking. The complementary pattern was observed in the subsample with underlying conditions or poor self-rated health, where abstainer scores were substantially elevated relative to the full sample. High-risk drinker scores remained substantially elevated across all three samples. A consistent pattern was observed for GAD-7 (Table 2).

### Sensitivity analysis

Sensitivity analyses using alternative lag specifications produced similar results, with a J-shaped association between sustained alcohol use and symptom scores in the full sample that was attenuated in the healthier subsample (Supplementary Table S4, Supplementary Figures S1-S2).

### Effect modification by sex

Interaction coefficients were small in magnitude and 95% bootstrap confidence intervals crossed zero for all three alcohol categories (low-risk: −0.29, 95% CI −1.73 to 1.38; moderate-risk: −0.11, 95% CI −1.92 to 1.84; high-risk: −0.21, 95% CI −3.36 to 3.06), providing no evidence of sex as an effect modifier (Supplementary Table S5).

## Discussion

In this longitudinal cohort study, we used inverse probability weighting and artificial censoring to estimate symptoms of anxiety and depression under sustained alcohol use strategies over a two-year period. Although the full sample showed a J-shaped association consistent with prior observational studies employing g-methods,^31,34^ stratified analyses suggest this pattern reflected residual confounding driven by sick-quitter bias rather than a protective effect of moderate drinking. Among participants without underlying conditions or poor self-rated health, differences across abstinent, low-risk, and moderate-risk drinking strategies were minimal, while high-risk drinking remained associated with higher symptom burden. In contrast, the J-shaped pattern was amplified among participants with underlying conditions or poor self-rated health, the group most susceptible to sick-quitter bias.^77^

Within-strategy comparisons further supported this interpretation. Attenuation in the healthier subsample was driven primarily by lower predicted symptom scores among abstainers and low-risk drinkers, while scores among moderate-risk drinkers were effectively unchanged. This suggests that the full-sample contrast was driven less by uniquely low symptom scores among moderate-risk drinkers than by elevated symptom scores among abstainers and low-risk drinkers with poorer health. As illustrated in Figure 2, analyses restricted to healthier participants reduce the likelihood of including individuals who reduced or stopped drinking because of declining health before cohort enrollment, when reasons for drinking reduction and the histories that could serve as proxies for sick-quitting were not available. In contrast, among participants with poorer health, prior health-related quitters may be present alongside other abstainers or low-risk drinkers, leaving greater potential for residual confounding. These findings align with evidence from studies of alcohol use, cardiovascular disease and mortality, where apparent benefits of moderate drinking relative to abstinence have been reinterpreted as artifacts of health-related selection into abstinence.^36^ Future work could more directly address sick-quitter confounding by collecting reasons for quitting or reducing drinking, longer drinking histories, and clinical measures of illness severity to better distinguish health-driven abstinence from other forms of abstinence and to identify which conditions most strongly contribute to sick-quitter bias.

The consistently elevated symptom burden among high-risk drinkers across all analyses reinforces the well-established bi-directional relationship between heavy alcohol use and anxiety and depression, and distinguishes this association from the apparent protective effect of moderate drinking, which did not persist after accounting for health-related selection. These findings support co-screening for unhealthy alcohol use alongside anxiety and depression and suggest that harm reduction approaches focused on reducing heavy drinking may serve as appropriate clinical targets for mental health promotion. Future research may evaluate whether these patterns replicate in samples with clinically diagnosed anxiety or depression, where self-medication pathways may be more pronounced and alcohol use may have more direct clinical implications for symptom trajectories. In such studies, more precise measurement of mental health treatment, including pharmacy or electronic health record data, would strengthen control for time-varying confounding by treatment type, dose, and consistency over time.

### Strengths and Contributions

This study has several strengths. The CHASING COVID Cohort provides a large, diverse U.S. sample with repeated assessments over three years. The use of artificial censoring and inverse probability weighting enabled estimation of outcomes under sustained alcohol-use strategies while accounting for time-varying confounding and loss to follow-up. Inclusion of a broad set of time-varying covariates extended confounding control beyond prior studies employing g-methods,^31,34^ while the persistence of the J-shaped pattern despite this approach highlights the challenges of fully addressing sick-quitter bias. In the absence of direct measures of reasons for quitting or reducing drinking, baseline health-status stratification provided an analytic strategy for evaluating whether the alcohol-symptom pattern changed in a manner consistent with health-related selection into abstinence.

### Limitations

Several limitations should be noted. Alcohol use and anxiety and depression symptoms were self-reported, introducing potential for misclassification. Generalizability may be limited because the cohort, while socio-demographically and geographically diverse, is not nationally representative. Our causal interpretation relies on assumptions of no unmeasured confounding, consistency, and positivity, which cannot be fully verified. For example, although we adjusted for many important confounders, personality characteristics such as extraversion, conscientiousness, and agreeableness were not measured and could contribute to residual confounding.^52^ In addition, the alcohol-use strategies examined here represent broad categories rather than fully specified interventions; individuals may arrive at or maintain the same category through different behavioral changes, and among those who drink, the same category may include different drinking patterns, which may have differing implications for mental health. Our estimates should therefore be interpreted as average effects over the distribution of alcohol-use patterns represented by each category in the study population, rather than effects of a single, precisely defined intervention.^78^ Furthermore, the stratification approach used to probe sick-quitter bias is informative but does not constitute a formal causal test, and residual differences between subgroups may persist; however, the consistent attenuation of the J-shaped association among healthier participants and its amplification among those with poorer health is difficult to explain by alternative mechanisms. Finally, the small number of female high-risk drinkers (n=164) limited our ability to detect sex-specific effects.

### Conclusions

These findings suggest that the observed J-shaped pattern between alcohol use and symptoms of anxiety and depression likely reflects sick-quitter bias rather than a protective effect of moderate drinking. The J-shaped pattern was concentrated among participants with underlying conditions or poor self-rated health and disappeared among healthier participants, while higher symptom scores under sustained high-risk drinking remained. These results underscore the need for caution in interpreting apparent benefits of moderate alcohol use and highlight both the value and limits of causal inference approaches in observational alcohol research.

## Acknowledgements

We thank the participants of the Communities, Households, and SARS-CoV-2 Epidemiology COVID Cohort Study for their contribution to the advancement of science.

## CRediT Author Statement

**Jenna Sanborn:** Conceptualization, Methodology, Software, Formal analysis, Investigation, Writing-Original Draft. **Denis Nash**: Writing-Review & Editing, Funding acquisition. **McKaylee Robertson:** Writing-Review & Editing. **Angela M Parcesepe**: Writing-Review & Editing, Funding acquisition. **Zachary Shahn:** Methodology, Writing-Review and Editing, Supervision.

## Data availability statement

The datasets generated and/or analyzed during the current study are available in the repository, Zenodo: <u>DOI: 10.5281/zenodo.6127734</u>. Some data elements are not publicly available due to funder requirements, but are available from the authors upon reasonable request, subject to approval and available resources.

