## Supplementary Material for "The Role of Sick-Quitter Bias in the Association Between Alcohol Use and Anxiety and Depression Symptoms: A Causal Analysis in a U.S. National Cohort"

### 1. ****Supplementary Methods: Statistical Analysis****

#### 1.1. Target trial specification

We emulated a hypothetical target trial to estimate the effect of sustained alcohol use strategies on anxiety and depression symptom severity. Eligible participants were adults in the CHASING COVID Cohort with valid alcohol consumption data at the September 2021 assessment, which served as time zero. Participants were followed through December 2023 across five subsequent assessment waves.

The interventions of interest were four sustained alcohol use strategies defined at each assessment: abstinent, low-risk, moderate-risk, and high/severe-risk drinking, based on AUDIT-C categories. The outcomes were anxiety and depression symptom severity at the final assessment (December 2023), measured using the GAD-7 and PHQ-8 scales and analyzed as continuous outcomes.

#### 1.2. Artificial censoring and adherence definition

To emulate sustained treatment strategies, we applied artificial censoring based on adherence to each strategy. At each timepoint, we defined a strategy-specific adherence indicator based on the participant’s observed alcohol consumption category. Participants were considered adherent if their AUDIT-C category matched the assigned strategy.

An “ever-adherent” indicator was constructed for each strategy using a cumulative product across timepoints, such that participants remained classified as adherent until the first deviation from the assigned strategy. For artificial censoring, person-time was retained through and including the timepoint at which the deviation occurred, with participants censored thereafter. Separately, participants were censored at the first timepoint with missing alcohol consumption data and at their last observed timepoint if outcome data were missing.

#### 1.3. Inverse probability weight estimation

We estimated stabilized inverse probability weights to account for both artificial censoring due to non-adherence and loss to follow-up. For each sustained alcohol use strategy, we fit pooled logistic regression models to estimate the probability of remaining adherent at each timepoint.

Denominator models included time (modeled as discrete assessment wave), baseline covariates, and lagged time-varying covariates measured at the two prior assessments and (t–1 and t–2). Numerator models included time and baseline covariates only. Predicted probabilities were truncated to the interval [1e-6, 1–1e-6] to avoid numerical instability and multiplied cumulatively across timepoints to generate individual-level stabilized adherence weights.

Separately, inverse probability of censoring weights were estimated to account for loss to follow-up using pooled logistic regression models with a similar structure. Final stabilized weights for each participant were calculated as the product of the adherence weight and the censoring weight.

#### 1.4. Weight diagnostics and truncation

We assessed the distribution of stabilized weights for each strategy using summary statistics, including the minimum, median, and upper percentiles, as well as effective sample size. For the abstinent and low-risk strategies, weights were well-behaved and were used without truncation. For the moderate- and high-risk strategies, extreme weights were observed, and we applied percentile-based truncation at the 1st and 99th percentiles to improve stability and reduce the influence of outliers.

#### 1.5. Outcome estimation and bootstrap inference

For each sustained alcohol use strategy, we estimated the weighted mean PHQ-8 and GAD-7 scores at the final assessment using the stabilized weights. Confidence intervals were obtained using nonparametric bootstrap resampling (B = 1000). In each bootstrap replicate, participants were resampled with replacement, and the full estimation procedure-including weight estimation and outcome calculation-was repeated. Point estimates were calculated as the mean of the bootstrap estimates, and 95% confidence intervals were derived from the 2.5th and 97.5th percentiles of the bootstrap distribution.

#### 1.6. Sensitivity analysis: alternative lag specification

As a sensitivity analysis, we repeated the full estimation procedure using an alternative lag structure for time-varying covariates. In this specification, covariates were drawn from the same survey wave as the exposure (t0) and the immediately preceding wave (t–1), instead of the primary approach using t–1 and t–2 lags. This approach may improve confounding control for time-sensitive variables by using more temporally proximal data, given that relying exclusively on longer lags (e.g., 3–6 months before exposure) increases the risk of residual confounding due to time-varying conditions that change rapidly. However, this specification assumes that covariates measured at t0 occurred prior to the exposure, and that these variables are more likely to influence drinking behavior during the same period rather than be influenced by it. Thus, to maintain strict temporal ordering, lag 1 and lag 2 were pre-specified for the primary analysis.

#### 1.7 Handling of missing data

Missingness across assessments is summarized in Supplementary Table S5. For time-varying covariates, missing values were handled using last observation carried forward (LOCF), whereby the most recent prior observed value was carried forward to subsequent timepoints. This approach was used to preserve temporal ordering and avoid excluding person-time due to intermittent missingness. Overall missingness across time-varying covariates ranged from 3.6%-9.7% by assessment.

Time-fixed measures for neighborhood community, life satisfaction, and spirituality/religion were first collected after enrollment (in February 2021) and missing 4.5% of values. These items were single-imputed given the low level of missingness (<5%) across all variables.

Participants were censored at the first timepoint at which alcohol consumption data were missing during follow-up. Participants missing outcome data at the final assessment (December 2023) were censored at their last observed timepoint. 10.1% of participants with valid baseline alcohol data were missing outcome data.

| **Supplementary Table S1. Definitions and assessment timing of variables used in analysis** | | | | |
| --- | --- | --- | --- | --- |
| Variable | Variable Type | Definition | Assessment Timing | Assessment Question(s) |
| Exposure & Outcomes | | | | |
| Alcohol consumption risk level* | Categorical | Calculated by summing scores from three assessment questions; Each has five answer choices, with point values ranging from 0 to 4. The final score ranges from 0 to 12, and is used to determine alcohol consumption risk level | Sept 2021, Dec 2021,  Mar 2022, Oct 2022, Apr 2023, Sept 2023 | In the past month, how often do you have a drink containing alcohol? [Never, Monthly or less, 2-4 times a month, 2-3 times a week, 4 or more times a week]  *If do not drink alcohol, then skip:* In the last month, how many standard drinks containing alcohol do you have on a typical day? [1 or 2, 3 or 4, 5 or 6, 7-9, 10 or more]  *If do not drink alcohol, then skip:* In the last month, how often do you have six or more drinks on one occasion? [Never, Less than monthly, Monthly, Weekly, Daily or almost daily] |
| GAD-7 score (anxiety)* | Continuous | Sum of scores where not at all =0, several days=1, over half the days=2 and nearly every day=3. The final score ranges from 0-21. | Dec 2023 | In the past month, how often have you been bothered by the following symptoms? [Not at all, Several days, Over half the days, Nearly every day]: 1) Feeling nervous, anxious or on edge, 2) Not being able to stop or control worrying, 3) Worrying too much about different things, 4) Trouble relaxing, 5) Being so restless it’s hard to sit still, 6) Becoming easily annoyed or irritable, 7) Feeling afraid as if something awful might happen |
| PHQ-8 score (depression)* | Continuous | Sum of scores, where not at all =0, several days=1, over half the days=2 and nearly every day=3. The final score ranges from 0-24. | Dec 2023 | In the past month, how often have you been bothered by the following symptoms? [Not at all, Several days, Over half the days, Nearly every day]: 1) Feeling down, depressed or hopeless, 2) Trouble falling or staying asleep, or sleeping too much, 3) Feeling tired or having little energy, 4) Poor appetite or overeating, 5) Feeling bad about yourself- or that you are a failure or have let yourself or your family down, 6) Trouble concentrating on things, such as, reading the newspaper or watching television, 7) Moving or speaking so slowly that other people have noticed? Or the opposite- being so fidgety or restless that you have been moving around a lot more than usual |
| Time-fixed covariates | | | | |
| Pre-baseline AUDIT-C scores | Continuous | Average of pre-baseline AUDIT-C scores | Enrollment,  Nov 2020,  May 2021 | In the past month, how often do you have a drink containing alcohol? [Never, Monthly or less, 2-4 times a month, 2-3 times a week, 4 or more times a week]  *If do not drink alcohol, then skip:* In the last month, how many standard drinks containing alcohol do you have on a typical day? [1 or 2, 3 or 4, 5 or 6, 7-9, 10 or more]  *If do not drink alcohol, then skip:* In the last month, how often do you have six or more drinks on one occasion? [Never, Less than monthly, Monthly, Weekly, Daily or almost daily] |
| Age | Continuous | Self-reported age | Enrollment | What is your age? Age ____ |
| Sex at birth | Binary | Collapsed to 2 categories: ‘Male’ and ‘Female’. Missing values, imputed with gender identity, assessed at baseline, if male or female was endorsed | April 2023 | What sex were you assigned at birth? a) Male, b) Female, c) None of the above |
| Race/ethnicity | Categorical | Self-reported race/ethnicity grouped into five categories: Hispanic, White (non-Hispanic), Black (non-Hispanic), Asian/ Pacific Islander (non-Hispanic), Other | Enrollment | Are you Hispanic, Latino/a, or Spanish origin? [Yes, No, Don’t know / Not sure]  Which of these groups would you say best represents your race? Please select all that apply. [Black or African American, American Indian or Alaska Native, Asian, Pacific Islander, White, Other ___, Don’t know/ Not sure] |
| Education | Categorical | Highest education level completed: [Less than high school, high school graduate, some college, college graduate] | Enrollment | What is the highest grade or year of school you completed? [Less than a high school diploma, Grade 12 or GED (High school graduate), College 1 year to 3 years (Some college or technical school), College 4 years or more (College graduate) |
| Annual Household Income | Categorical | Annual household income from all sources: [<$35,000, $50,000-$69,999, $70,000-99,999, $100,000+] | Enrollment | Is your annual household income from all sources: [<$25,000, $25,000-$34,999, $35,000-$49,999, $50,000-$69,999, $70,000-$99,999, $100,000-$149,000, $150,000+] |
| Body Mass Index | Categorical | Calculated as kg/m^2^ based on height and weight and categorized as Underweight, Healthy weight, Overweight, Obesity [ref] | Enrollment | How much do you weigh without shoes? Please answer in pounds ____  About how tall are you without shoes? Please answer in feet and inches. ____ Feet ____ Inches |
| Cigarette smoking status | Binary | Coded as ‘1’ if some days or every day selected | Enrollment | Do you currently smoke cigarettes every day, some days or not at all? (Cigarettes does not include electronic products such as: e-cigarettes, vape pens, personal vaporizers, e-cigars, epipes, e-hookahs, hookah pens, and mods). a) Every day, b) Some days, c) Not at all, d) Don’t know / Not sure |
| Household size | Categorical | Sum total of number of people in each age group (children, 18-59 and 60+) | Enrollment | How many members of your household, including yourself, are between 18-59 years of age? [___number, no one aged 18-59]  How many members of your household, including yourself, are 60 years old or older? [___number, no one aged 60+]  How many children less than 18 years of age live in your household? [___number, no children < 18 live in my household] |
| Residential area type | Categorical | Residential area designation based on zip code categorized as “Suburban/Town”, ‘Rural” and “Urban” | Enrollment | ZIP code was used to create the geographic-level variable for residential area type, assigned based on the NCES Education Demographic and Geographic Estimates locale definitions, using the ZCTA locale file to map ZIP codes to 'Rural', 'Suburban', 'Urban', and 'Town'. Given the low number of 'Town' designations (n=7), ‘Suburban’ and 'Town’ were collapsed into a single category. |
| Neighborhood community | Categorical | Sense of neighborhood community [Strongly agree/agree, Neutral, Stronger disagree/disagree] | Feb 2021 | Living in my current neighborhood gives me a strong sense of community. [Strongly agree, Agree, Neutral, Disagree, Strongly Disagree] |
| Life satisfaction | Continuous | Level of satisfaction with life as a whole. | Feb 2021 | Overall, how satisfied are you with life as a whole these days? (0= not satisfied at all, 10= completely satisfied) |
| Spirituality/ Religion | Categorical | Faith, religion, spirituality gives sense of purpose [Strongly agree/agree, Neutral, Stronger disagree/disagree] | Feb 2021 | My faith, religious, or spiritual beliefs give me a sense of direction and purpose in my life. [Strongly agree, Agree, Neutral, Disagree, Strongly disagree] |
| Time-Varying Covariates | | | | |
| GAD-7 score  (anxiety) | Continuous | Sum of scores where not at all =0, several days=1, over half the days=2 and nearly every day=3. The final score ranges from 0-21. | Pre-baseline (avg from assessments between enrollment and May 2020), Sept 2021, Dec 2021, Mar 2022, Jun 2022, Oct 2022, Dec 2022, Apr 2023, Jun 2023, Sept 2023 | In the past month, how often have you been bothered by the following symptoms? [Not at all, Several days, Over half the days, Nearly every day]: 1) Feeling nervous, anxious or on edge, 2) Not being able to stop or control worrying, 3) Worrying too much about different things, 4) Trouble relaxing, 5) Being so restless it’s hard to sit still, 6) Becoming easily annoyed or irritable, 7) Feeling afraid as if something awful might happen |
| PHQ-8 score (depression) | Continuous | Sum of scores, where not at all =0, several days=1, over half the days=2 and nearly every day=3. The final score ranges from 0-24. | Pre-baseline (avg from assessments between July 2020 and May 2020), Sept 2021, Dec 2021, Mar 2022, Jun 2022, Oct 2022, Dec 2022, Apr 2023, Jun 2023, Sept 2023 | In the past month, how often have you been bothered by the following symptoms? [Not at all, Several days, Over half the days, Nearly every day]: 1) Feeling down, depressed or hopeless, 2) Trouble falling or staying asleep, or sleeping too much, 3) Feeling tired or having little energy, 4) Poor appetite or overeating, 5) Feeling bad about yourself- or that you are a failure or have let yourself or your family down, 6) Trouble concentrating on things, such as, reading the newspaper or watching television, 7) Moving or speaking so slowly that other people have noticed? Or the opposite- being so fidgety or restless that you have been moving around a lot more than usual |
| Physical activity | Continuous | Minutes of physical activity per week calculated using the three assessment questions | Oct 2020, Feb 2021, May 2021, Sept 2021, Dec 2021, Mar 2022, Jun 2022, Oct 2022, Dec 2022, Apr 2023, Jun 2023, Sept 2023 | 1. During the past month, other than your regular job, did you participate in any physical activities or exercises such as running, calisthenics, golf, gardening or walking for exercise? [Yes, No, Don’t know/ Not sure]  2. How many times per week or per month did you take part in this activity during the past month? [__ times per week, __ times per month, don’t know/ not sure]  3. And when you took part in this activity, for how many minutes or hours did you usually keep at it? [__ Number of hours, __ Number of minutes, Don’t know/ Not sure] |
| Mental health treatment | Categorical | Received counseling from a mental health professional in the previous 4 weeks [Yes vs. No/don’t know]  Or  Took prescription medication for mental health in the previous 4 weeks  Categorized as neither, both, counseling only, Rx only | May 2021, Sept 2021, Dec 2021, Mar 2022, Oct 2022, Apr 2023, Sept 2023 | In the past four weeks, have you received counseling or therapy from a mental health professional, such as a psychologist, social worker, or other licensed mental health professional? [Yes, No, Don’t know/ Not sure]  In the past four weeks, have you taken prescription medication for your mental health? [Yes, No, Don’t know/ Not sure] |
| Mental health diagnosis | Binary | Diagnosed with anxiety, depression or PTSD (yes vs. no) | Enrollment,  Dec 2021, Sept 2023 | Has a doctor, nurse, or other health professional ever told you that you had any of the following? *Select all that apply* [Anxiety, depression, PTSD] |
| Health insurance status | Binary | Participants categorized as having insurance vs. not (yes vs. no/don’t know). | Enrollment, Feb 2021, May 2021,  Sept 2021, Dec 2021,  Mar 2022, Jun 2022,  Oct 2022, Dec 2022,  Apr 2023, Jun 2023,  Sept 2023 | Do you have any kind of health care coverage, including health insurance, prepaid plans such as HMOs, or government plans such as Medicare, or Indian Health Service? [Yes, No, Don’t know/ Not sure] |
| Housing instability | Binary | Categorize as housing insecure if “always” or “usually” is selected | Enrollment, Feb 2021, May 2021, Sept 2021, Dec 2021, Mar 2022, Oct 2022, Apr 2023, Sept 2023 | How often in the past month would you say you were worried or stressed about having enough money to pay your rent/mortgage? [Always, Usually, Sometimes, Rarely, Never] |
| Food insecurity | Binary | Categorize as food insecure if “Often” or “Sometimes” is selected for either of these statements^148^ | Enrollment, Feb 2021, May 2021,  Sept 2021, Dec 2021,  Mar 2022, Oct 2022, Apr 2023, Sept 2023 | For each of the following, you will answer whether the statement was often true, sometimes true, or never true for (you/your household) in the past month.  1) “We worried whether our food would run out before we got money to buy more.” [Often true, sometimes true, never true]  2) “The food that we bought just didn’t last, and we didn’t have money to get more.” Was that often, sometimes or never true for you in the past month? [Often true, sometimes true, never true] |
| Income loss | Binary | Categorized as having lost income vs. not (‘Yes’ vs. ‘No/ not applicable’) | Enrollment, Feb 2021, May 2021, Sept 2021, Dec 2021, Mar 2022, Jun 2022, Oct 2022, Dec 2022, Apr 2023 | In the past month, have you experienced a significant personal loss of income as a result of COVID-19? [Yes, no, not applicable] |
| Employment status | Categorical | Responses classified into three categories: [Employed, Unemployed, Not in labor force] | Enrollment, Feb 2021, May 2021, Sept 2021, Dec 2021, Mar 2022, Jun 2022, Oct 2022, Dec 2022, Apr 2023, Jun 2023, Sept 2023 | Are you currently…? [Employed for wages, Self-employed, Out of work for less than 1 year, Out of work for 1 year or more, A homemaker, A student, Retired] |
| General health status | Categorical | General health categories: [Poor, Fair, Very Good, Excellent] | July 2020, Feb 2021, May 2021, Sept 2021, Dec 2021, Mar 2022, Jun 2022, Oct 2022, Dec 2022, Apr 2023, Jun 2023, Sept 2023 | Would you say that in general your health is: [Excellent, Very Good, Fair, Poor] |
| Number of chronic conditions | Numeric | Sum total of ever diagnosed with the following health conditions:  *diabetes, high blood pressure, chronic lung disease or COPD, heart attack, angina, kidney disease, immunocompromised condition, HIV, or current asthma* | Enrollment, Dec 2021, Sept 2023 | Has a doctor, nurse, or other health professional ever told you that you had any of the following? *Select all that apply*  had a heart attack also called a myocardial infarction? b) have angina or coronary heart disease? c) have type 2 diabetes? d) have high blood pressure? e) have cancer? f) had asthma? g) have chronic obstructive pulmonary disease, C.O.P.D., emphysema or chronic bronchitis? h) have kidney disease (not including kidney stones, bladder infection or incontinence)? i) have HIV/AIDS? j) have immunosuppression? k) have depression? l) have post-traumatic stress disorder or PTSD? m) have chronic liver disease, including cirrhosis? n) have an anxiety disorder? o) I have not been told that I have any of the above conditions  If yes to asthma: Do you still have asthma? [Yes, No, Don’t know/ Not sure] |
| Poor physical health days | Continuous | Number of days as a continuous number | July 2020, Feb 2021, May 2021, Sept 2021, Dec 2021, Mar 2022, Jun 2022, Oct 2022, Dec 2022, Apr 2023, Jun 2023, Sept 2023 | Now thinking about your physical health, which includes physical illness and injury, for how many days during the past 30 days was your physical health not good? a) _____ Number of days from 1-30 b) None c) Don’t know / Not sure |
| Poor mental health days | Continuous | Number of days as a continuous number | July 2020, Feb 2021, May 2021, Sept 2021, Dec 2021, Mar 2022, Jun 2022, Oct 2022, Dec 2022, Apr 2023, Jun 2023, Sept 2023 | Now thinking about your mental health, which includes stress, depression, and problems with emotions, for how many days during the past 30 days was your mental health not good? a) _____ Number of days from 1-30 b) None c) Don’t know / Not sure |
| Disability status | Categorical | Responses classified into three categories: [No difficulty, Some difficulty, A lot of difficulty] | July 2020, Feb 2021, May 2021, Sept 2021, Dec 2021, Mar 2022, Jun 2022, Oct 2022, Dec 2022, Apr 2023, Jun 2023, Sept 2023 | Since your last survey (ADD Qualtrics DD/Mon/YY), how much difficulty do you have engaging in daily activities (or household responsibilities) because of physical, mental, or emotional problems? a) No difficulty b) Some difficulty c) A lot of difficulty d) Don’t know / Not sure |
| Social engagement level | Categorical | Calculated number of social activities participants spent time doing during the past month: 1) Gathering of 10+ people, either indoors or outdoors, 2) Spent time inside of a house that is not your own, 3) Spent time inside a restaurant or bar, 4) Spent time in the patio or outdoor space of a restaurant or bar, or 5) Spent time at any of the following: The inside of a restaurant or bar, A patio or outdoor space at a restaurant or bar, An indoor movie theatre, A shopping mall, A church, synagogue, mosque or other place of worship, The inside of a house that is not your own, an overnight stay at the residence of family or friends.  Responses categorized as ‘low’, ‘medium’ and ‘high’: low= <2, medium: 2-4, high: 5+ | July 2020, Feb 2021, May 2021, Sept 2021, Dec 2021, Mar 2022 | In the past month, have you gathered in groups with 10 or more people? a) Yes, indoors only, b) Yes, outdoors only, c) Yes, indoors and outdoors, d) No, e) Don’t know/ Not sure  In the past month, have you done any of the following? [Yes, No, Not applicable]  a) Spent time inside of a house that is not your own, b) Spent time inside a restaurant or bar, c) Spent time in the patio or outdoor space of a restaurant or bar, d) Had an overnight stay at a hotel, short-term rental or residence of family or friends  In the past month, have you spent time in any of the following places? Please select all that apply.  a) A hairdresser, salon or barber, b) The inside of a restaurant or bar, c) A patio or outdoor space at a restaurant or bar, d) An indoor movie theatre, e) A shopping mall, f) A church, synagogue, mosque or other place of worship, g) The inside of a house that is not your own, h) A public swimming area such as a pool, lake, ocean or bay, i) a public park, j) a mass gathering like a demonstration or public protest, k) A mass gathering like a political rally, l) a hotel or other short-term rental (like Airbnb) where people outside of your household are staying, m) An overnight stay at the residence of family or friends, n) An overnight trip to another town or city, o) A gym or exercise facility, p) None of the above |
| Drug use | Binary | Responses collapsed to two categories: “Never”, “Once or twice”, vs. ”Daily” or “Almost daily” | Dec 2020, May 2021, Sept 2021, Dec 2021,  Mar 2022, Oct 2022, Apr 2023, Sept 2023 | In the past month how many times have you used the following: [Never, Once or twice, Weekly, Daily or almost daily]: 1) Cannabis (marijuana, pot, grass, hash, etc.), 2) Street opioids (heroin, opium, etc.), 3) Prescription opioids in a way or dose other than prescribed (fentanyl, oxycodone, hydrocodone, methadone, buprenorphine, etc.) |
| Substance use or alcohol recovery | Binary | Responses collapsed to two categories: “Yes- currently or previously” vs. “Never/ Not sure” | Enrollment, May 2021, Sept 2021, Dec 2021,  Mar 2022, Oct 2022, Apr 2023, Sept 2023 | Do you identify as being in recovery from drugs? [Yes, I am currently in recovery from drugs, I am not currently in recovery from drugs, but I have previously been in recovery from drugs, I have never been in recovery from drugs, Don’t know/ Not sure]  Do you identify as being in recovery from alcohol? [Yes, I am currently in recovery from alcohol, I am not currently in recovery from alcohol, but I have previously been in recovery from alcohol, I have never been in recovery from alcohol, Don’t know/ Not sure] |
| Receives government food support | Binary | Classify responses into two categories: 'Any government programs’ vs. ‘none’ | Feb 2021, Dec 2021, Mar 2022, Oct 2022, Apr 2023, Sept 2023 | In the past month, have you used any of the following? *Select all that apply*. [None, food pantry, soup kitchen, SNAP, pEBT, emergency food support, other] |
| Relationship status | Binary | Classify responses into two categories: ‘Yes’ vs. ‘No/ Don’t know’ | Enrollment, May 2021, Sept 2021, Dec 2021, Apr 2023, Sept 2023 | Are you currently in a relationship or seeing someone? [Yes, No, Don’t know/ Not sure] |
| COVID-19 Vaccination status | Binary | Binary variable indicating whether two doses of the vaccine had been received as of the date of the survey assessment. | Feb 2021, May 2021, Sept 2021, Dec 2021, Mar 2022, Jun 2022, Oct 2022, Dec 2022, Apr 2023, Jun 2023, Sept 2023 | Have you been fully or partially vaccinated against COVID-19 with a vaccine that has received FDA approval or emergency use authorization? [Yes, No, Don’t know / Not sure]  If yes to fully or partially vaccinated in this survey: How many doses of the primary vaccine series did you receive? Primary vaccine series means either a 2-dose mRNA COVID-19 vaccine series (Moderna or Pfizer) or a single dose of Johnson & Johnson COVID-19 vaccine. If you received booster doses please do not include them here. [1, 2]  If received 1 dose only or 2 doses: When did you receive your first dose of the COVID-19 vaccine? Your vaccination card should have the date of your first shot. [Enter date: Month Day Year lookup]  If received 2 doses: When did you receive your second dose of the COVID-19 vaccine? Your vaccination card should have the date of your second shot. [Enter date: Month Day Year lookup] |

| **Supplementary Table S2. Participant characteristics by analytic subsample, CHASING COVID Cohort, September 2021** | | | |
| --- | --- | --- | --- |
|  | **Full sample** | **Subsample without underlying conditions/ poor health** | **Subsample with underlying conditions/ poor health** |
| **Characteristic** | **N = 4,673**^1^ | **N = 2,794**^1^ | **N = 1,860**^1^ |
| Anxiety Score (GAD-7) | 5.36 (5.33) | 5.04 (5.10) | 5.83 (5.65) |
| Depression Score (PHQ-8) | 5.82 (5.83) | 5.27 (5.51) | 6.64 (6.20) |
| Alcohol consumption category (AUDIT-C) |  |  |  |
| Abstinence | 1,418 (30.3%) | 769 (27.5%) | 642 (34.5%) |
| Low-risk | 1,748 (37.4%) | 1,056 (37.8%) | 685 (36.8%) |
| Moderate-risk | 1,069 (22.9%) | 709 (25.4%) | 357 (19.2%) |
| High-risk | 438 (9.4%) | 260 (9.3%) | 176 (9.5%) |
| Age (years) | 42.72 (15.09) | 39.50 (13.91) | 47.59 (15.51) |
| Sex |  |  |  |
| Male | 2,117 (45.3%) | 1,193 (42.7%) | 918 (49.4%) |
| Female | 2,556 (54.7%) | 1,601 (57.3%) | 942 (50.6%) |
| Race/Ethnicity |  |  |  |
| Hispanic | 749 (16.0%) | 472 (16.9%) | 272 (14.6%) |
| White (non-Hispanic) | 2,973 (63.6%) | 1,741 (62.3%) | 1,224 (65.8%) |
| Black (non-Hispanic) | 447 (9.6%) | 229 (8.2%) | 214 (11.5%) |
| Asian/Pacific Islander (non-Hispanic) | 342 (7.3%) | 268 (9.6%) | 73 (3.9%) |
| Other (non-Hispanic) | 162 (3.5%) | 84 (3.0%) | 77 (4.1%) |
| Education |  |  |  |
| Less than high school | 68 (1.5%) | 32 (1.1%) | 35 (1.9%) |
| High school | 446 (9.5%) | 232 (8.3%) | 211 (11.3%) |
| Some college | 1,180 (25.3%) | 640 (22.9%) | 534 (28.7%) |
| College graduate | 2,979 (63.7%) | 1,890 (67.6%) | 1,080 (58.1%) |
| Household size |  |  |  |
| 1 | 1,180 (25.3%) | 653 (23.4%) | 523 (28.1%) |
| 2 | 1,528 (32.7%) | 891 (31.9%) | 633 (34.0%) |
| 3 | 731 (15.6%) | 445 (15.9%) | 282 (15.2%) |
| 4 | 661 (14.1%) | 436 (15.6%) | 223 (12.0%) |
| 5+ | 573 (12.3%) | 369 (13.2%) | 199 (10.7%) |
| Residential area type |  |  |  |
| Suburban/Town | 1,228 (26.3%) | 727 (26.0%) | 496 (26.7%) |
| Rural | 1,400 (30.0%) | 774 (27.7%) | 621 (33.4%) |
| Urban | 2,045 (43.8%) | 1,293 (46.3%) | 743 (39.9%) |
| Annual household income |  |  |  |
| <$35,000 | 1,252 (26.8%) | 662 (23.7%) | 583 (31.3%) |
| $35,000–$49,999 | 537 (11.5%) | 300 (10.7%) | 233 (12.5%) |
| $50,000–$69,999 | 713 (15.3%) | 419 (15.0%) | 291 (15.6%) |
| $70,000–$99,000 | 804 (17.2%) | 496 (17.8%) | 308 (16.6%) |
| $100,000+ | 1,367 (29.3%) | 917 (32.8%) | 445 (23.9%) |
| Employment status |  |  |  |
| Employed | 3,172 (67.9%) | 2,062 (73.8%) | 1,100 (59.1%) |
| Unemployed | 383 (8.2%) | 176 (6.3%) | 204 (11.0%) |
| Not in labor force | 1,118 (23.9%) | 556 (19.9%) | 556 (29.9%) |
| Lost income in previous month |  |  |  |
| No | 4,096 (87.7%) | 2,512 (89.9%) | 1,568 (84.3%) |
| Yes | 577 (12.3%) | 282 (10.1%) | 292 (15.7%) |
| Has health insurance |  |  |  |
| No | 464 (9.9%) | 293 (10.5%) | 166 (8.9%) |
| Yes | 4,209 (90.1%) | 2,501 (89.5%) | 1,694 (91.1%) |
| Housing insecurity |  |  |  |
| No | 3,399 (72.7%) | 2,122 (75.9%) | 1,270 (68.3%) |
| Yes | 1,274 (27.3%) | 672 (24.1%) | 590 (31.7%) |
| Food insecurity |  |  |  |
| No | 3,761 (80.5%) | 2,357 (84.4%) | 1,394 (74.9%) |
| Yes | 912 (19.5%) | 437 (15.6%) | 466 (25.1%) |
| Received government food assistance |  |  |  |
| No | 3,514 (75.2%) | 2,209 (79.1%) | 1,297 (69.7%) |
| Yes | 1,159 (24.8%) | 585 (20.9%) | 563 (30.3%) |
| Physically active (60+ mins per week) |  |  |  |
| No | 1,828 (39.1%) | 968 (34.6%) | 849 (45.6%) |
| Yes | 2,845 (60.9%) | 1,826 (65.4%) | 1,011 (54.4%) |
| BMI category |  |  |  |
| Underweight | 99 (2.1%) | 72 (2.6%) | 27 (1.5%) |
| Healthy weight | 1,744 (37.3%) | 1,285 (46.0%) | 453 (24.4%) |
| Overweight | 1,411 (30.2%) | 850 (30.4%) | 554 (29.8%) |
| Obesity | 1,419 (30.4%) | 587 (21.0%) | 826 (44.4%) |
| Has chronic health condition |  |  |  |
| No | 2,881 (61.7%) | 2,794 (100.0%) | 68 (3.7%) |
| Yes | 1,792 (38.3%) | 0 (0.0%) | 1,792 (96.3%) |
| Daily smoker |  |  |  |
| No | 4,041 (86.5%) | 2,493 (89.2%) | 1,536 (82.6%) |
| Yes | 632 (13.5%) | 301 (10.8%) | 324 (17.4%) |
| General health status |  |  |  |
| Excellent | 107 (2.3%) | 7 (0.3%) | 100 (5.4%) |
| Good | 1,178 (25.2%) | 492 (17.6%) | 681 (36.6%) |
| Fair | 2,489 (53.3%) | 1,596 (57.1%) | 882 (47.4%) |
| Poor | 899 (19.2%) | 699 (25.0%) | 197 (10.6%) |
| Vaccinated for COVID-19 |  |  |  |
| No | 660 (14.1%) | 417 (14.9%) | 242 (13.0%) |
| Yes | 4,013 (85.9%) | 2,377 (85.1%) | 1,618 (87.0%) |
| Received mental health diagnosis |  |  |  |
| No | 789 (16.9%) | 484 (17.3%) | 303 (16.3%) |
| Yes | 3,884 (83.1%) | 2,310 (82.7%) | 1,557 (83.7%) |
| Received mental health treatment |  |  |  |
| No | 3,081 (81.6%) | 1,946 (85.8%) | 1,124 (75.3%) |
| Yes | 693 (18.4%) | 321 (14.2%) | 369 (24.7%) |
| Used any drugs in the past month |  |  |  |
| No | 3,358 (71.9%) | 2,022 (72.4%) | 1,324 (71.2%) |
| Yes | 1,315 (28.1%) | 772 (27.6%) | 536 (28.8%) |
| Identifies as being in recovery for drugs and/or alcohol |  |  |  |
| No | 4,054 (86.8%) | 2,483 (88.9%) | 1,556 (83.7%) |
| Yes | 619 (13.2%) | 311 (11.1%) | 304 (16.3%) |
| Relationship status |  |  |  |
| Not in a relationship | 1,752 (37.5%) | 966 (34.6%) | 779 (41.9%) |
| In a relationship | 2,921 (62.5%) | 1,828 (65.4%) | 1,081 (58.1%) |
| Life satisfaction |  |  |  |
| Low | 626 (13.4%) | 333 (11.9%) | 290 (15.6%) |
| Medium | 1,552 (33.2%) | 920 (32.9%) | 624 (33.5%) |
| High | 2,495 (53.4%) | 1,541 (55.2%) | 946 (50.9%) |
| Social engagement level |  |  |  |
| Low | 1,066 (22.8%) | 555 (19.9%) | 502 (27.0%) |
| Medium | 2,286 (48.9%) | 1,376 (49.2%) | 903 (48.5%) |
| High | 1,321 (28.3%) | 863 (30.9%) | 455 (24.5%) |
| **Strong sense of neighborhood community** |  |  |  |
| Strongly agree | 517 (11.1%) | 293 (10.5%) | 220 (11.8%) |
| Agree | 1,408 (30.1%) | 832 (29.8%) | 573 (30.8%) |
| Neutral | 1,780 (38.1%) | 1,104 (39.5%) | 670 (36.0%) |
| Disagree | 674 (14.4%) | 397 (14.2%) | 274 (14.7%) |
| Strongly disagree | 294 (6.3%) | 168 (6.0%) | 123 (6.6%) |
| **Faith, religion or spirituality gives sense of purpose** |  |  |  |
| Strongly agree | 801 (25.4%) | 446 (24.3%) | 352 (27.0%) |
| Agree | 1,060 (33.6%) | 604 (32.9%) | 452 (34.6%) |
| Neutral | 815 (25.8%) | 479 (26.1%) | 330 (25.3%) |
| Disagree | 240 (7.6%) | 155 (8.4%) | 85 (6.5%) |
| Strongly disagree | 239 (7.6%) | 152 (8.3%) | 86 (6.6%) |
| ^1^Mean (SD); n (%) | | | |

Supplementary Table S3. Participant characteristics by baseline alcohol consumption among adults without underlying conditions or poor general health, CHASING COVID Cohort, September 2021 (N=2,813)**

|  | **Alcohol consumption levels** | | | | |  |
| --- | --- | --- | --- | --- | --- | --- |
| **Characteristic** | Overall^1^ | **Abstinent**  N = 776^1^ | **Low risk**  N = 1,063^1^ | **Moderate risk**  N = 712^1^ | **High/severe risk**  N = 262^1^ | **p-value**^2^ |
| **Anxiety Score (GAD-7)** | 5.04 (5.09) | 5.28 (5.58) | 4.76 (4.96) | 4.63 (4.38) | 6.59 (5.60) | <0.001 |
| **Depression Score (PHQ-8)** | 5.28 (5.51) | 5.51 (6.00) | 4.97 (5.37) | 4.81 (4.79) | 7.18 (5.93) | <0.001 |
| **Age (years)** | 39.50 (13.91) | 38.77 (14.58) | 39.78 (13.42) | 40.76 (14.71) | 37.08 (10.87) | 0.004 |
| **Sex** |  |  |  |  |  | <0.001 |
| Male | 1,199 (42.6%) | 285 (36.7%) | 520 (48.9%) | 232 (32.6%) | 162 (61.8%) |  |
| Female | 1,614 (57.4%) | 491 (63.3%) | 543 (51.1%) | 480 (67.4%) | 100 (38.2%) |  |
| **Race/Ethnicity** |  |  |  |  |  | <0.001 |
| Hispanic | 477 (17.0%) | 160 (20.6%) | 179 (16.8%) | 89 (12.5%) | 49 (18.7%) |  |
| White (non-Hispanic) | 1,749 (62.2%) | 411 (53.0%) | 650 (61.1%) | 518 (72.8%) | 170 (64.9%) |  |
| Black (non-Hispanic) | 233 (8.3%) | 80 (10.3%) | 91 (8.6%) | 35 (4.9%) | 27 (10.3%) |  |
| Asian/Pacific Islander (non-Hispanic) | 269 (9.6%) | 99 (12.8%) | 108 (10.2%) | 51 (7.2%) | 11 (4.2%) |  |
| Other (non-Hispanic) | 85 (3.0%) | 26 (3.4%) | 35 (3.3%) | 19 (2.7%) | 5 (1.9%) |  |
| **Education** |  |  |  |  |  | <0.001 |
| Less than high school | 33 (1.2%) | 20 (2.6%) | 5 (0.5%) | 7 (1.0%) | 1 (0.4%) |  |
| High school | 235 (8.4%) | 101 (13.0%) | 75 (7.1%) | 37 (5.2%) | 22 (8.4%) |  |
| Some college | 646 (23.0%) | 223 (28.7%) | 238 (22.4%) | 113 (15.9%) | 72 (27.5%) |  |
| College graduate | 1,899 (67.5%) | 432 (55.7%) | 745 (70.1%) | 555 (77.9%) | 167 (63.7%) |  |
| **Household size** |  |  |  |  |  | <0.001 |
| 1 | 657 (23.4%) | 170 (21.9%) | 267 (25.1%) | 155 (21.8%) | 65 (24.8%) |  |
| 2 | 895 (31.8%) | 215 (27.7%) | 334 (31.4%) | 257 (36.1%) | 89 (34.0%) |  |
| 3 | 449 (16.0%) | 127 (16.4%) | 159 (15.0%) | 114 (16.0%) | 49 (18.7%) |  |
| 4 | 438 (15.6%) | 122 (15.7%) | 184 (17.3%) | 98 (13.8%) | 34 (13.0%) |  |
| 5+ | 374 (13.3%) | 142 (18.3%) | 119 (11.2%) | 88 (12.4%) | 25 (9.5%) |  |
| **Residential area type** |  |  |  |  |  | <0.001 |
| Suburban/Town | 732 (26.0%) | 208 (26.8%) | 293 (27.6%) | 180 (25.3%) | 51 (19.5%) |  |
| Rural | 779 (27.7%) | 253 (32.6%) | 296 (27.8%) | 172 (24.2%) | 58 (22.1%) |  |
| Urban | 1,302 (46.3%) | 315 (40.6%) | 474 (44.6%) | 360 (50.6%) | 153 (58.4%) |  |
| **Annual household income** |  |  |  |  |  | <0.001 |
| <$35,000 | 669 (23.8%) | 252 (32.5%) | 249 (23.4%) | 111 (15.6%) | 57 (21.8%) |  |
| $35,000–$49,999 | 304 (10.8%) | 103 (13.3%) | 113 (10.6%) | 60 (8.4%) | 28 (10.7%) |  |
| $50,000–$69,999 | 422 (15.0%) | 126 (16.2%) | 164 (15.4%) | 94 (13.2%) | 38 (14.5%) |  |
| $70,000–$99,000 | 496 (17.6%) | 115 (14.8%) | 189 (17.8%) | 148 (20.8%) | 44 (16.8%) |  |
| $100,000+ | 922 (32.8%) | 180 (23.2%) | 348 (32.7%) | 299 (42.0%) | 95 (36.3%) |  |
| **Employment status** |  |  |  |  |  | <0.001 |
| Employed | 2,072 (73.7%) | 493 (63.5%) | 826 (77.7%) | 545 (76.5%) | 208 (79.4%) |  |
| Unemployed | 179 (6.4%) | 73 (9.4%) | 51 (4.8%) | 32 (4.5%) | 23 (8.8%) |  |
| Not in labor force | 562 (20.0%) | 210 (27.1%) | 186 (17.5%) | 135 (19.0%) | 31 (11.8%) |  |
| **Lost income in previous month** |  |  |  |  |  | <0.001 |
| No | 2,528 (89.9%) | 679 (87.5%) | 971 (91.3%) | 659 (92.6%) | 219 (83.6%) |  |
| Yes | 285 (10.1%) | 97 (12.5%) | 92 (8.7%) | 53 (7.4%) | 43 (16.4%) |  |
| **Has health insurance** |  |  |  |  |  | <0.001 |
| No | 298 (10.6%) | 127 (16.4%) | 86 (8.1%) | 48 (6.7%) | 37 (14.1%) |  |
| Yes | 2,515 (89.4%) | 649 (83.6%) | 977 (91.9%) | 664 (93.3%) | 225 (85.9%) |  |
| **Housing insecurity** |  |  |  |  |  | <0.001 |
| No | 2,129 (75.7%) | 547 (70.5%) | 833 (78.4%) | 580 (81.5%) | 169 (64.5%) |  |
| Yes | 684 (24.3%) | 229 (29.5%) | 230 (21.6%) | 132 (18.5%) | 93 (35.5%) |  |
| **Food insecurity** |  |  |  |  |  | <0.001 |
| No | 2,367 (84.1%) | 609 (78.5%) | 923 (86.8%) | 639 (89.7%) | 196 (74.8%) |  |
| Yes | 446 (15.9%) | 167 (21.5%) | 140 (13.2%) | 73 (10.3%) | 66 (25.2%) |  |
| **Received government food assistance** |  |  |  |  |  | <0.001 |
| No | 2,217 (78.8%) | 558 (71.9%) | 858 (80.7%) | 609 (85.5%) | 192 (73.3%) |  |
| Yes | 596 (21.2%) | 218 (28.1%) | 205 (19.3%) | 103 (14.5%) | 70 (26.7%) |  |
| **Physically active (60+ mins per week)** |  |  |  |  |  | <0.001 |
| No | 979 (34.8%) | 359 (46.3%) | 353 (33.2%) | 170 (23.9%) | 97 (37.0%) |  |
| Yes | 1,834 (65.2%) | 417 (53.7%) | 710 (66.8%) | 542 (76.1%) | 165 (63.0%) |  |
| **BMI category** |  |  |  |  |  | <0.001 |
| Underweight | 72 (2.6%) | 35 (4.5%) | 23 (2.2%) | 10 (1.4%) | 4 (1.5%) |  |
| Healthy weight | 1,291 (45.9%) | 352 (45.4%) | 455 (42.8%) | 369 (51.8%) | 115 (43.9%) |  |
| Overweight | 857 (30.5%) | 225 (29.0%) | 321 (30.2%) | 219 (30.8%) | 92 (35.1%) |  |
| Obesity | 593 (21.1%) | 164 (21.1%) | 264 (24.8%) | 114 (16.0%) | 51 (19.5%) |  |
| **Has chronic health condition** |  |  |  |  |  |  |
| No | 2,813 (100.0%) | 776 (100.0%) | 1,063 (100.0%) | 712 (100.0%) | 262 (100.0%) |  |
| Yes | 0 (0.0%) | 0 (0.0%) | 0 (0.0%) | 0 (0.0%) | 0 (0.0%) |  |
| **Daily smoker** |  |  |  |  |  | <0.001 |
| No | 2,505 (89.1%) | 666 (85.8%) | 972 (91.4%) | 659 (92.6%) | 208 (79.4%) |  |
| Yes | 308 (10.9%) | 110 (14.2%) | 91 (8.6%) | 53 (7.4%) | 54 (20.6%) |  |
| **General health status** |  |  |  |  |  | <0.001 |
| Excellent | 7 (0.2%) | 2 (0.3%) | 3 (0.3%) | 1 (0.1%) | 1 (0.4%) |  |
| Good | 497 (17.7%) | 192 (24.7%) | 159 (15.0%) | 92 (12.9%) | 54 (20.6%) |  |
| Fair | 1,607 (57.1%) | 410 (52.8%) | 648 (61.0%) | 409 (57.4%) | 140 (53.4%) |  |
| Poor | 702 (25.0%) | 172 (22.2%) | 253 (23.8%) | 210 (29.5%) | 67 (25.6%) |  |
| **Vaccinated for COVID-19** |  |  |  |  |  | <0.001 |
| No | 418 (14.9%) | 182 (23.5%) | 131 (12.3%) | 58 (8.1%) | 47 (17.9%) |  |
| Yes | 2,395 (85.1%) | 594 (76.5%) | 932 (87.7%) | 654 (91.9%) | 215 (82.1%) |  |
| **Received mental health diagnosis** |  |  |  |  |  | <0.001 |
| No | 486 (17.3%) | 84 (10.8%) | 196 (18.4%) | 158 (22.2%) | 48 (18.3%) |  |
| Yes | 2,327 (82.7%) | 692 (89.2%) | 867 (81.6%) | 554 (77.8%) | 214 (81.7%) |  |
| **Received mental health treatment** |  |  |  |  |  | 0.113 |
| No | 1,957 (85.8%) | 548 (88.5%) | 743 (84.6%) | 482 (85.6%) | 184 (83.3%) |  |
| Yes | 324 (14.2%) | 71 (11.5%) | 135 (15.4%) | 81 (14.4%) | 37 (16.7%) |  |
| **Used any drugs in the past month** |  |  |  |  |  | <0.001 |
| No | 2,034 (72.3%) | 649 (83.6%) | 805 (75.7%) | 452 (63.5%) | 128 (48.9%) |  |
| Yes | 779 (27.7%) | 127 (16.4%) | 258 (24.3%) | 260 (36.5%) | 134 (51.1%) |  |
| **Identifies as being in recovery for drugs and/or alcohol** |  |  |  |  |  | <0.001 |
| No | 2,498 (88.8%) | 629 (81.1%) | 1,001 (94.2%) | 657 (92.3%) | 211 (80.5%) |  |
| Yes | 315 (11.2%) | 147 (18.9%) | 62 (5.8%) | 55 (7.7%) | 51 (19.5%) |  |
| **Relationship status** |  |  |  |  |  | <0.001 |
| Not in a relationship | 973 (34.6%) | 338 (43.6%) | 347 (32.6%) | 196 (27.5%) | 92 (35.1%) |  |
| In a relationship | 1,840 (65.4%) | 438 (56.4%) | 716 (67.4%) | 516 (72.5%) | 170 (64.9%) |  |
| **Life satisfaction** |  |  |  |  |  | 0.002 |
| Low | 336 (11.9%) | 104 (13.4%) | 119 (11.2%) | 71 (10.0%) | 42 (16.0%) |  |
| Medium | 928 (33.0%) | 244 (31.4%) | 330 (31.0%) | 251 (35.3%) | 103 (39.3%) |  |
| High | 1,549 (55.1%) | 428 (55.2%) | 614 (57.8%) | 390 (54.8%) | 117 (44.7%) |  |
| **Social engagement level** |  |  |  |  |  | <0.001 |
| Low | 564 (20.0%) | 296 (38.1%) | 152 (14.3%) | 73 (10.3%) | 43 (16.4%) |  |
| Medium | 1,383 (49.2%) | 349 (45.0%) | 547 (51.5%) | 369 (51.8%) | 118 (45.0%) |  |
| High | 866 (30.8%) | 131 (16.9%) | 364 (34.2%) | 270 (37.9%) | 101 (38.5%) |  |
| **Strong sense of neighborhood community** |  |  |  |  |  | 0.239 |
| Strongly agree | 297 (10.6%) | 94 (12.1%) | 107 (10.1%) | 70 (9.8%) | 26 (9.9%) |  |
| Agree | 835 (29.7%) | 208 (26.8%) | 319 (30.0%) | 223 (31.3%) | 85 (32.4%) |  |
| Neutral | 1,110 (39.5%) | 315 (40.6%) | 424 (39.9%) | 276 (38.8%) | 95 (36.3%) |  |
| Disagree | 400 (14.2%) | 102 (13.1%) | 153 (14.4%) | 110 (15.4%) | 35 (13.4%) |  |
| Strongly disagree | 171 (6.1%) | 57 (7.3%) | 60 (5.6%) | 33 (4.6%) | 21 (8.0%) |  |
| **Faith, religion or spirituality gives sense of purpose** |  |  |  |  |  | <0.001 |
| Strongly agree | 449 (24.3%) | 191 (33.7%) | 148 (20.9%) | 72 (17.4%) | 38 (23.5%) |  |
| Agree | 608 (32.9%) | 168 (29.6%) | 256 (36.2%) | 142 (34.3%) | 42 (25.9%) |  |
| Neutral | 485 (26.2%) | 138 (24.3%) | 188 (26.6%) | 115 (27.8%) | 44 (27.2%) |  |
| Disagree | 155 (8.4%) | 30 (5.3%) | 64 (9.1%) | 47 (11.4%) | 14 (8.6%) |  |
| Strongly disagree | 153 (8.3%) | 40 (7.1%) | 51 (7.2%) | 38 (9.2%) | 24 (14.8%) |  |
| ^1^Mean (SD); n (%) | | | | | | |
| ^2^Kruskal-Wallis rank sum test; Pearson's Chi-squared test; NA | | | | | | |

| **Supplementary Table S4. Predicted Mean GAD-7 and PHQ-8 scores by Alcohol Use Trajectory (Sensitivity analysis with alternative lag specifications)**  *(Point Estimates and 95% Confidence Intervals)* | | | | |
| --- | --- | --- | --- | --- |
|  | GAD-7 Full Cohort (N=4,673) | GAD-7 Subsample  (N=2,813) | PHQ-8 Full Cohort  (N=4,673) | PHQ-8 Subsample (N=2,813) |
| Abstinent | 3.99 [3.63, 4.38] | 3.36 [2.93, 3.93] | 4.75 [4.34, 5.21] | 3.63 [3.17, 4.28] |
| Low risk | 3.24 [2.91, 3.65] | 3.04 [2.61, 3.53] | 3.99 [3.56, 4.52] | 3.42 [3.01, 3.89] |
| Moderate risk | 3.01 [2.48, 3.63] | 3.32 [2.76, 4.09] | 3.32 [2.80, 3.95] | 3.33 [2.72, 4.18] |
| High risk | 5.23 [4.24, 6.68] | 4.57 [3.37, 6.15] | 5.80 [4.54, 7.33] | 5.18 [3.92, 6.61] |
| Abstinent – Moderate contrast | 0.98 [0.26, 1.64] | 0.05 [-0.82, 0.84] | 1.43 [0.69, 2.15] | 0.30 [-0.62, 1.20] |
| **Subsample is restricted to participants without underlying conditions or poor general health* | | | | |

**Supplementary Figure S1. Predicted Mean PHQ-8 Scores Under Sustained Alcohol Use Trajectories
(Sensitivity analysis with alternative lag specification)**

**Fig. S1A. Full Cohort	Fig. S1B. Healthy Stratum**

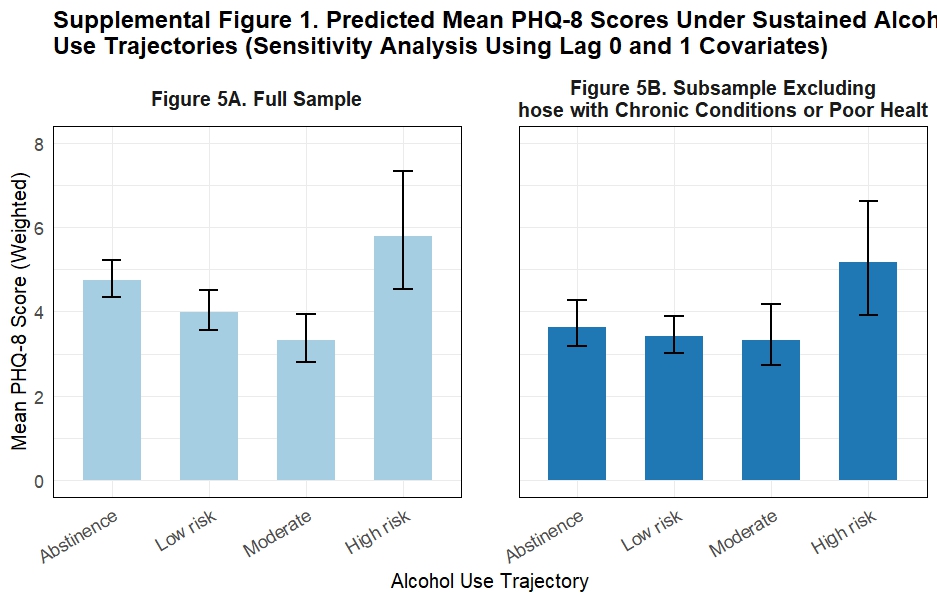

**Subsample is restricted to participants without underlying conditions or poor general health*

**Supplementary Figure S2. Predicted Mean GAD-7 Scores Under Sustained Alcohol Use Trajectories
(Sensitivity analysis with alternative lag specification)**

**Fig. S2A. Full Cohort	Fig. S2B. Healthy Stratum**

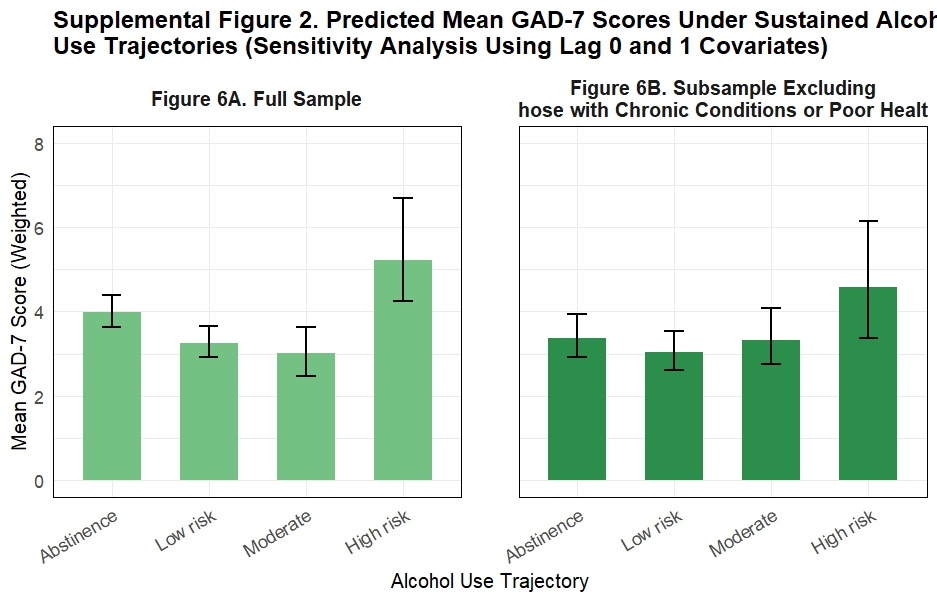

**Supplementary Figures S1-S2. Predicted Mean Mental Health Scores Under Sustained Alcohol Use Trajectories (Sensitivity Analysis Using Lag 0 and 1 Covariates). Figure 1** presents predicted mean PHQ-8 scores by alcohol use trajectory in the full sample (S1A) and a subsample excluding participants with chronic conditions or poor general health (S1B), using covariates measured at the same and one timepoint prior to the outcome while Supplementary Figure S2 shows corresponding predicted GAD-7 scores (S2A–B). Findings from the revised analysis using lag 0 and lag 1 covariates were largely consistent with the primary analysis (lag 1 and lag 2). Predicted mean GAD-7 and PHQ-8 scores across alcohol use trajectories were nearly identical, indicating that using covariates measured earlier in time did not materially change the observed associations. These results suggest that confounding by contemporaneous factors was minimal and that the primary findings were robust to alternative lag specifications.

| **Supplemental Table S5. Test of Effect Modification by Sex: Interaction Between Sex and Sustained Alcohol Use Category on Mean Depression (PHQ-8) and Anxiety (GAD-7) Symptom Severity, CHASING COVID Cohort, September 2021-December 2023** | | | | |
| --- | --- | --- | --- | --- |
| **Alcohol use category (vs. abstinent)** | **PHQ-8** | | **GAD-7** | |
|  | **Coefficient (95% CI)^†^** | **p-value** | **Coefficient (95% CI)^†^** | **p-value** |
| Full cohort (N=4,673) |  |  |  |  |
| Low-risk | -0.53 (-2.03, 1.03) | 0.49 | -0.46 (-1.73, 0.87) | 0.46 |
| Moderate-risk | -0.29 (-1.80, 1.36) | 0.76 | 0.19 (-1.17, 1.60) | 0.75 |
| High/severe-risk | -0.88 (-3.59, 2.36) | 0.63 | -1.02 (-3.47, 1.87) | 0.53 |
| Healthy Stratum^*^ (2,813) |  |  |  |  |
| Low-risk | −0.29 (-1.73 to 1.38) | 0.72 | -0.70 (-2.14 to 0.75) | 0.40 |
| Moderate-risk | −0.11 (-1.92 to 1.84) | 0.96 | 0.35 (-1.41 to 2.16) | 0.65 |
| High/severe-risk | −0.21 (-3.36 to 3.06 | 0.80 | -0.33 (-3.44 to 2.38) | 0.76 |
| *^†^Interaction coefficient estimated from a weighted linear regression of the outcome on alcohol use category, sex, and their product terms, using the same stabilized inverse-probability weights applied in the primary analysis. Males are the reference category. The coefficient represents the difference in the alcohol-vs.-abstinent symptom score contrast for females relative to males. A negative value indicates the contrast is smaller (less positive) for females than for males. *Subsample is restricted to participants without underlying conditions or poor general health CI = confidence interval; GAD-7 = Generalized Anxiety Disorder-7; PHQ-8 = Patient Health Questionnaire-8. Bootstrap p-values and confidence intervals derived from 1,000 bootstrap replicates.* | | | | |

Supplementary Table S6. Missing data patterns across

assessments (September 2021–December 2023), CHASING

COVID Cohort (N=4,673)

| Number of missing surveys | N | % |
| --- | --- | --- |
| 0 | 3,724 | 72.3 |
| 1 | 472 | 9.2 |
| 2 | 194 | 3.8 |
| 3 | 140 | 2.7 |
| 4 | 134 | 2.6 |
| 5 | 119 | 2.3 |
| 6 | 98 | 1.9 |
| 7 | 92 | 1.8 |
| 8 | 102 | 2.0 |
| 9 | 77 | 1.5 |
